# Hospital unit colonization pressure and nosocomial acquisition of drug susceptible and drug resistant pathogens

**DOI:** 10.1101/2025.06.11.25329430

**Authors:** Luke Sagers, Ziming Wei, Caroline McKenna, Christina Chan, Anna A. Agan, Theodore R. Pak, Chanu Rhee, Michael Klompas, Sanjat Kanjilal

## Abstract

**Background:** Hospitalized patients are at risk for developing hospital acquired infections. Active surveillance for bacterial colonization is effective at preventing infection but is resource-intensive and limited to high-risk units and a subset of high-risk pathogens. Colonization pressure (CP) for common pathogens has been associated with hospital acquired infection and can be calculated in real-time using data in the electronic health record. We aimed to assess the impact of CP on nosocomial acquisition for a range of drug susceptible and drug resistant pathogens, across an entire hospital system.

**Methods:** We conducted a retrospective matched cohort study of all inpatients admitted to a large regional integrated healthcare system between May 2015 and July 2024 who stayed in one room during the 30 day observation period. Cases had target organisms detected in any clinical or surveillance culture taken between 3 and 30 days after admission into their first room. Controls were matched on demographics, length of stay, prior surgery and 14 classes of antibiotic exposure. Our outcome was nosocomial acquisition of 11 common pathogens spread across enteric, skin and environmental niches. We applied conditional logistic regression and XGBoost to model nosocomial acquisition using as covariates the Elixhauser comorbidity index and CP, defined as the time-weighted prevalence of an organism in ward co-occupants over the previous 6 months. CP was calculated for 9 organism sets, including the Enterobacterales, ESBL Enterobacterales, vancomycin susceptible and resistant *Enterococcus* spp, *C. difficile*, methicillin susceptible *Staphylococcus aureus* (MSSA), methicillin resistant *S. aureus* (MRSA) and drug susceptible and drug resistant (DR) *P. aeruginosa* (PsA).

**Findings:** Our pooled cohort included 14,923 cases matched to 28,480 controls. Hospital acquisition occurred four times more frequently for drug susceptible versus drug resistant organisms. Baseline characteristics were well matched between cases and controls. The strongest positive associations were between CP*_C. difficile_* and nosocomial acquisition of *C. difficile* (+32.5%, 95% CI +21.9% to +44.0%), CP_ESBL_ and ESBL *K. pneumoniae* (+29.4%, 95% CI +11.3% to +50.6%), and CP_PsA-DR_ with drug resistant *P. aeruginosa* (+28.6%, 95% CI +14.0% to +45.0%). Among the skin flora, CP_MSSA_ was associated with a +12.1% (95% CI +9.9% to +14.4%) increase in the odds of nosocomial acquisition of MSSA, CP_MRSA_ was associated with a +6.7% (95% CI +1.3% to +12.5%) increase in the odds of MRSA. Negative associations were observed between organisms inhabiting different niches, including CP_MSSA_ and ESBL *K. pneumoniae* (-7.9%, 95% CI -15.1% to -0.2%), and CP_PsA-DR_ and vancomycin susceptible *E. faecalis* (-10.0%, 95% CI -15.6% to -4.0%).

**Interpretation:** A hospitalized patient’s odds of nosocomially acquiring a potential pathogen is associated with its prevalence among that patient’s ward co-occupants, regardless of the organism’s drug resistance profile. Further research is necessary to understand the role of passive surveillance of CP for preventing infection.

**Funding:** LWS was supported by the NLM (2T15LM007092-31). ZW was supported with institutional funding from the Department of Population Medicine. TRP was supported by AHRQ (K08-HS030118). SK was supported by AHRQ (grant no. K08 HS027841-01A1).

## INTRODUCTION

Hospital acquired infection is a common and serious complication of medical care^1^. A primary mechanism for this begins when a patient who is colonized with a potential pathogen is admitted to the hospital. That individual becomes a reservoir from which the hands and clothing of healthcare workers, hospital equipment and room surfaces are contaminated^2^. Transfer from these contaminated surfaces, often via the hands of healthcare workers^3^, results in transmission to and colonization of a new vulnerable host^4^. Colonization is a strong predictor of future clinical infection^5–7^. The cycle repeats when the second patient becomes a new reservoir for onward nosocomial transmission.

Active surveillance for colonization of asymptomatic individuals is a key component of infection control programs, but requires significant investment in infrastructure and human resources. For this reason it is typically limited to intensive care units (ICUs) and other high-risk units, and to a few drug resistant or high-virulence organisms, such as methicillin resistant *Staphylococcus aureus* (MRSA) and vancomycin resistant *Enterococcus* species (VRE). Colonization pressure (CP), defined as the prevalence of an organism among patients in the ward into which a patient enters, has previously been shown to be associated with hospital acquired infection^8–10^. CP surveillance has the potential to augment active surveillance efforts as its estimation relies solely on information already present in the electronic health record (EHR). Additionally, it can be calculated for any number of organisms, in any area of the hospital, whereas expanding active surveillance can be disruptive to care and costly.

Prior analyses examining the impact of CP on hospital acquired infection were limited to known high-risk clonal pathogens and to ICU settings^10–13^. Whether the same relationship applies to drug susceptible organisms, which are at least as common as drug-resistant organisms^14,15^, and to non-ICU settings, remains unknown. Thus, the objectives of this study are twofold. The first is to build a prototype of an open source infection control informatics tool that takes raw EHR data and constructs hospital unit-level CP across a variety of common organisms and to publish a de-identified patient level CP dataset for reproducibility and generalizability. Second, we use our our prototype to test the hypothesis that CP is associated with hospital acquisition for a wide range of drug susceptible and drug resistant organisms relative to controls matched by demographics, length of stay, surgery, and fine-grained antibiotic exposures.

## METHODS

This study was deemed exempt by the Institutional Review Board of Mass General Brigham. We have adhered to STROBE and RECORD reporting guidelines.

### Study design

We conducted a matched retrospective cohort study using data obtained from the Mass General Brigham (MGB) healthcare system’s EHR data warehouse. MGB includes 8 community and 2 tertiary care hospitals in the New England area. Patients were included in the study if they were adults (>=18 years old) admitted to any MGB hospital between May 25 2015 and July 7 2024.

### Cohort definition

A case was defined as a patient with a clinical or surveillance specimen obtained from any body site with the target organism during the observation period, which was set as day +3 to day +30 after entry into their inpatient room. Controls were matched 2:1 by age ±5 years, sex, and history of any surgery in the previous 90 days. Controls were also required to have a length of stay in their room that was within ±3 days of a case’s length of stay, or the date timestamp of collection for the case’s index culture, whichever was shorter. Finally, controls were matched on the number of courses of antibiotics (inpatient and outpatient) received in the previous 60 days. A course was defined as >2 days of treatment >7 days apart and was stratified by the following antibiotic classes: penicillins, anti-staphylococcal penicillins, extended-spectrum penicillins, cephalosporins, extended-spectrum cephalosporins, carbapenems, glycopeptides, fluoroquinolones, macrolides, tetracyclines, anti-anaerobic antibiotics and a separate category for oral vancomycin for targeting *C. difficile*. A full list of antibiotics within each treatment class category is located in supplementary table S1.

Both cases and controls were required to have stayed in only one room for >48 hours during the entirety of the observation period. Collection of the index culture after departure from the room was allowed if the patient had no other stay for >48 hours between day +3 to day +30. Other study inclusion criteria included not being actively treated for infection, defined as no record of antibiotic exposure from day -7 to day +3 relative to the time of entry into their room. Both cases and controls also had to have no prior evidence of the organism of interest in the previous 12 months for *Staphylococcus aureus* and *Enterococcus* species, and the previous 6 months for all other target organisms. Figure 1 depicts the study design.

**Figure 1:**
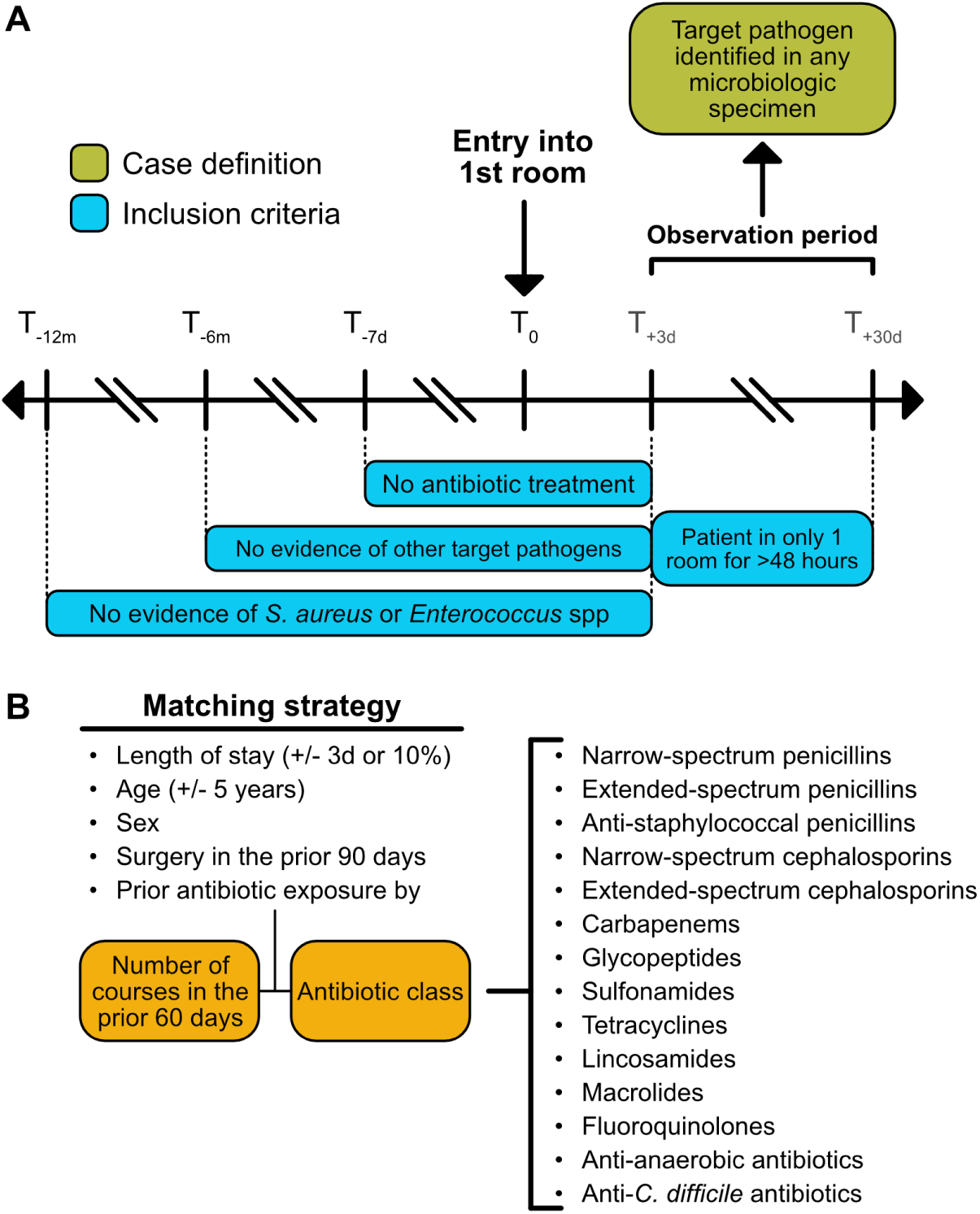
Study design. A) T_0_ was set as the time stamp of entry into the room for cases and controls. This room always represented the first room in a patient’s hospital stay that lasted >48 hours, not including the emergency room. All study participants were observed for culture positivity with the target organism between day +3 to day +30 after entry into their first room. All study participants had no evidence of antibiotic exposure in the previous 7 days, nor any clinical or surveillance cultures with a target organism in the previous 6 months (12 months if the target organism was *S. aureus* or an *Enterococcus* species). They also had a single room stay for >48 hours during the observation period. B) Matching strategy of cases to controls. Cases were matched to controls based on age, sex, evidence of surgery in the previous 90 days, and antibiotic exposure. The latter was further stratified by the number of courses consisting of ≥2 days of exposure that were >7 days apart, in the previous 60 days, across 14 drug classes.

### Target organisms

We built cohorts to predict nosocomial acquisition for 11 different organisms across skin, enteric and environmental niches. Among enteric organisms, we selected drug susceptible and extended-spectrum beta-lactamase (ESBL) producing *E. coli* and *Klebsiella pneumoniae*, vancomycin susceptible *E. faecalis*, vancomycin resistant *E. faecium*, and *C. difficile*. A drug susceptible *E. coli* and *K. pneumoniae* phenotype was defined as an organism susceptible to 3rd and 4th generation cephalosporins, and piperacillin-tazobactam, while the ESBL-producing phenotype was defined as resistance to any 3rd or 4th generation cephalosporin or piperacillin-tazobactam. Both drug susceptible and ESBL Enterobacterales phenotypes retained susceptibility to carbapenems. Among skin organisms, we selected methicillin susceptible *S. aureus* (MSSA) and MRSA. Among environmental organisms, we selected drug susceptible *Pseudomonas aeruginosa*, defined as an organism susceptible to ceftazidime, cefepime and piperacillin-tazobactam, and drug resistant *P. aeruginosa*, defined as resistance to any one of the anti-Pseudomonal β-lactams.

### Model covariates

We utilized colonization pressure (CP) to estimate the impact of the hospital environment on the odds of a hospital acquisition. We defined CP as the time-weighted prevalence of a specified organism, or organism set (as in the Enterobacterales), in ward co-occupants, given by:

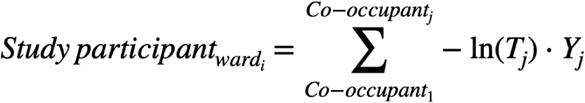

where *Y_j_* = 1 if the ward co-occupant had the target organism within the past 6 months (*Y_j_* = 0 if not) and *T_j_* is the time interval in days between the date the organism was identified in the co-occupant and the date of entry into the ward by the study participant (case or control). A ward co-occupant was defined as a person with a >48 hour stay in the 30-day period prior to entry into the ward by the study participant. The look-back period for *Y_j_* and *T_j_*was set to 6 months to account for the possibility of organism carriage, and the value was set to 0.001 if the co-occupant never had the organism. A negative log transform was applied to *T_j_* to account for the diminishing effect of prior organisms over time. CP was calculated separately for the following 9 organisms and groups: drug susceptible Enterobacterales and ESBL Enterobacterales, vancomycin susceptible (VSE) and vancomycin resistant *Enterococcus* (VRE) species, *C. difficile*, MSSA, MRSA, and drug susceptible (DS-PsA) and drug resistant *P. aeruginosa* (DR-PsA). Drug susceptible, drug resistant, and ESBL phenotypes are defined as above for the target organisms. Figure 2 illustrates the calculation of CP. Models also included patient comorbidities, encoded by the Agency for Healthcare Quality Elixhauser Mortality Index^16^.

**Figure 2:**
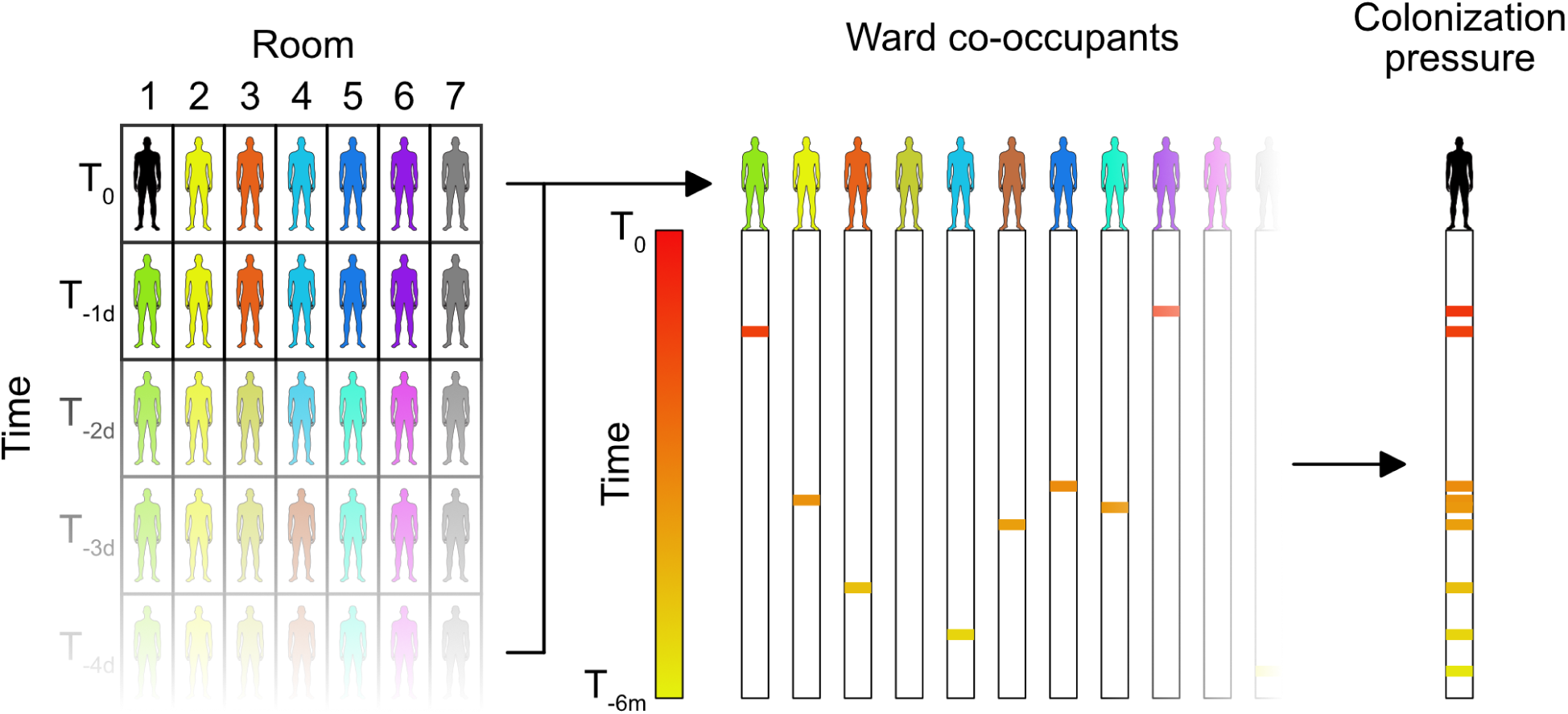
Definition of colonization pressure. Colonization pressure (CP) was defined as the time-weighted prevalence of a specified organism, or organism sets, in ward co-occupants. In the figure, the case or control enters Room 1 at T_0_. CP was calculated by identifying ward co-occupants in the previous 30 days and recording the number of days since they had the CP organism (or organism set) in the previous 6 months. A negative log transform was then applied to the number of days to account for diminishing effect over time and values were summed across all ward co-occupants to calculate the final value.

### Statistical analyses

We utilized conditional logistic regression for estimating the impact of organism-specific CP on the odds of nosocomial acquisition, given the matched case-control design and controlling for all other CP organisms and comorbidity burden. 95% confidence intervals were determined using the Wald method, as implemented in the survival R package. Statistical significance was determined based on whether the confidence interval around exponentiated coefficients crossed 1. The performance of the conditional logistic regression models was evaluated using a 5-fold cross-validation. In each fold, 20% of the groups within the matched dataset were used as the validation set and removed from training. The area under the receiver operator characteristic (AUROC) for the conditional logistic regression models was based on performance in the validation set. 95% confidence intervals for AUROCs were calculated using the standard deviation across the five bootstrapped validation folds. We report results separately for cognate relationships, defined as the target organism and the CP organism/s belonging to the same niche (e.g. skin, enteric, or environmental), and non-cognate relationships, where they inhabit different niches.

XGBoost models^17^ using the same covariates were used to further verify the result of the conditional logistic regression. The performance of XGBoost models was also evaluated through 5-fold cross-validation. In each fold, the full dataset was divided into training, validation, and test sets, with a ratio of 8:1:1. The performance was evaluated on the held-out test set in each fold, and the 95% confidence interval of the AUROC was estimated using the standard deviation of AUROC across the five folds.

A grid search was used to determine the optimum hyperparameters of the conditional logistic regression and XGBoost models. Feature importance was determined using the exponentiated coefficient of individual features for the conditional logistic regression and with Shapley values^18^ for XGB models. All analyses were performed using R (version 4.4.0) and Python (version 3.6.15). The source code for the entire analytic pipeline, spanning pre-processing of raw data to model output, is available on <u>Github</u>^19^, and a de-identified version of the final cleaned dataset used for the CP models is available on Physionet^20^ (project title ‘Nosocomial pathogen acquisition due to hospital unit colonization pressure’).

## RESULTS

### Baseline characteristics

A total of 18,626 patients met inclusion criteria, with 14,923 cases of hospital acquired organisms matched to 28,480 controls. Sample sizes for individual target organisms ranged from 610 for vancomycin resistant *E. faecium* (244 cases matched to 366 controls), to 9,460 for MSSA (2,956 cases matched to 6,504 controls). Nosocomial acquisition occurred nearly 4 times more frequently for drug susceptible organisms (*E. coli*, *K. pneumoniae*, vancomycin susceptible *E. faecalis*, MSSA, drug susceptible *P. aeruginosa*), than for drug resistant organisms (ESBL *E. coli*, ESBL *K. pneumoniae*, vancomycin resistant *E. faecium*, MRSA, drug resistant *P. aeruginosa*). Flow diagrams for individual cohorts are shown in supplementary table 2.

Baseline characteristics were similar across cases and controls for each target organism. Mean age for cases was 63.6 years (SE ± 0.2 years) and 63.9 years (SE ± 0.1 years) for controls. Ages ranged from 52.1 years (SE ± 0.5 years), and 54.7 years (SE ± 0.3 years) for MSSA cases and controls, respectively, to 70.7 years (SE ± 0.9 years), and 70.7 years (SE ± 0.8 years) for vancomycin resistant *E. faecium* cases and controls, respectively. Similarly, mean Elixhauser index ranged from 8.8 (SE ± 0.3) and 7.3 (SE ± 0.2) for MSSA case and controls, respectively, to 14.3 (SE ± 1.0), and 12.0 (SE ± 0.8) for vancomycin resistant *E. faecium* cases and controls, respectively. The number of prior antibiotic courses per 100 persons ranged from 3.5 and 2.8 for *E. coli* cases and controls, respectively to 9.0 and 7.7 for *C. difficile* cases and controls, respectively (Figure 3). Baseline features for the pooled cohort are shown in Table 1. Relevant characteristics for individual organism cohorts are shown in Figure 3, and supplementary tables S3 to S7. The distribution of comorbidities by individual Elixhauser categories, class-level antibiotic exposures, and the cumulative distribution of colonization pressures are shown in supplementary figures S1 to S5.

**Figure 3:**
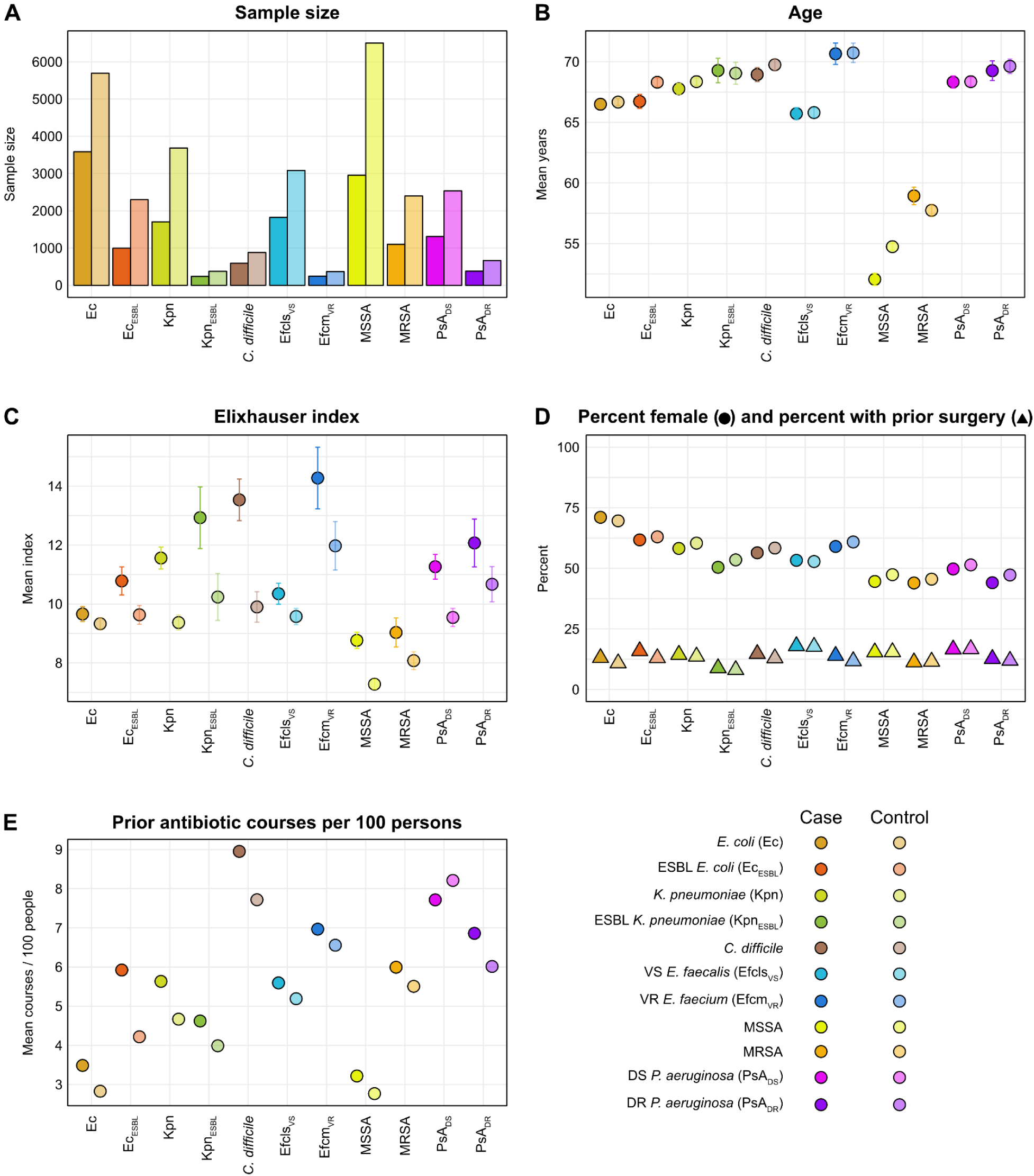
Select baseline characteristics across 11 cohorts of target organisms. Error bars represent standard errors of the mean. Cases and controls were matched by age, sex, length of stay, history of any surgery in the previous 90 days and number of antibiotic courses by class. Models included the Elixhauser Mortality Index. Abbreviations, VS, vancomycin susceptible; VR, vancomycin resistant; DS, drug susceptible; DR drug resistant.

**Table 1:** Baseline characteristics in pooled cohort across all organisms.

| Feature | Pooled cohort |  |
| --- | --- | --- |
|  | Cases | Controls |
| Sample size | 14,923 | 28,480 |
| Demographics |  |  |
| Age (mean, SE) | 63.6 +/- 0.2 | 63.9 +/- 0.1 |
| Gender (% female) | 55.9 | 56.1 |
| Elixhauser index (mean, SE) | 10.3 +/- 0.1 | 8.9 +/- 0.1 |
| Prior surgery (%) | 14.6 | 13.8 |
| Length of Stay (median, IQR) | 5.0 (3.2, 9.0) | 3.9 (2.9, 5.5) |
| Matching Duration (median, IQR) | 4.1 (3.1, 5.9) | 3.9 (2.9, 5.5) |
| Time to Infection (median, IQR) | 8.8 (5.0, 17.1) | -- |
| Prior antimicrobials (%) |  |  |
| β-lactams |  |  |
| Penicillins | 0.2 | 0.2 |
| Extended spectrum penicillins | 0.7 | 0.6 |
| Cephalosporins | 1.2 | 1.1 |
| Extended spectrum cephalosporins | 0.0 | 0.0 |
| Anti-Staphylococcal β-lactams | 0.0 | 0.0 |
| Other cell wall active agents |  |  |
| Glycopeptides | 0.0 | 0.0 |
| DNA synthesis inhibitors |  |  |
| Sulfonamides | 0.8 | 0.7 |
| Protein synthesis inhibitors |  |  |
| Fluoroquinolones | 1.1 | 1.0 |
| Tetracyclines | 0.6 | 0.5 |
| Macrolides | 0.4 | 0.4 |
| Lincosamides | 0.1 | 0.1 |
| Anti-anaerobic antibiotics |  |  |
| Metronidazole | 0.1 | 0.1 |
| Oral vancomycin | 0.0 | 0.0 |

### Colonization pressure and nosocomial acquisition among cognate pairs

The adjusted odds of hospital acquisition of an organism due to cognate colonization pressures is shown in Figure 4. Among the enteric flora, the strongest positive association was between CP*_C. difficile_* and hospital acquisition of *C. difficile* (+32.5%, 95% CI +21.9% to +44.0%). Similarly, CP_VRE_ and CP_ESBL_ were both associated with an increase in the odds of ESBL *K. pneumoniae*, at +29.4% (95% CI +11.3% to +50.6%), and +10.4% (95% CI +2.6% to +18.8%), respectively. A unit increase in CP_VSE_ was associated with an increase in the odds of five enteric organisms: ESBL *E. coli* (+5.0%, 95% CI +1.3% to +8.9%), drug susceptible *K. pneumoniae* (+2.9%, 95% CI 0.0% to +5.9%), *C. difficile* (+6.2%, 95% CI +0.4% to +12.2%), vancomycin susceptible *E. faecalis* (+11.8%, 95% CI 8.6% to 15.2%), and vancomycin resistant *E. faecium* (+8.6%, 95% CI +0.8% to +17.1%). Finally, a unit increase in CP_Enterobacterales_ was associated with a +1.0% (95% CI +0.2% to +1.8%) increase in the odds of acquiring drug susceptible *E. coli*.

**Figure 4:**
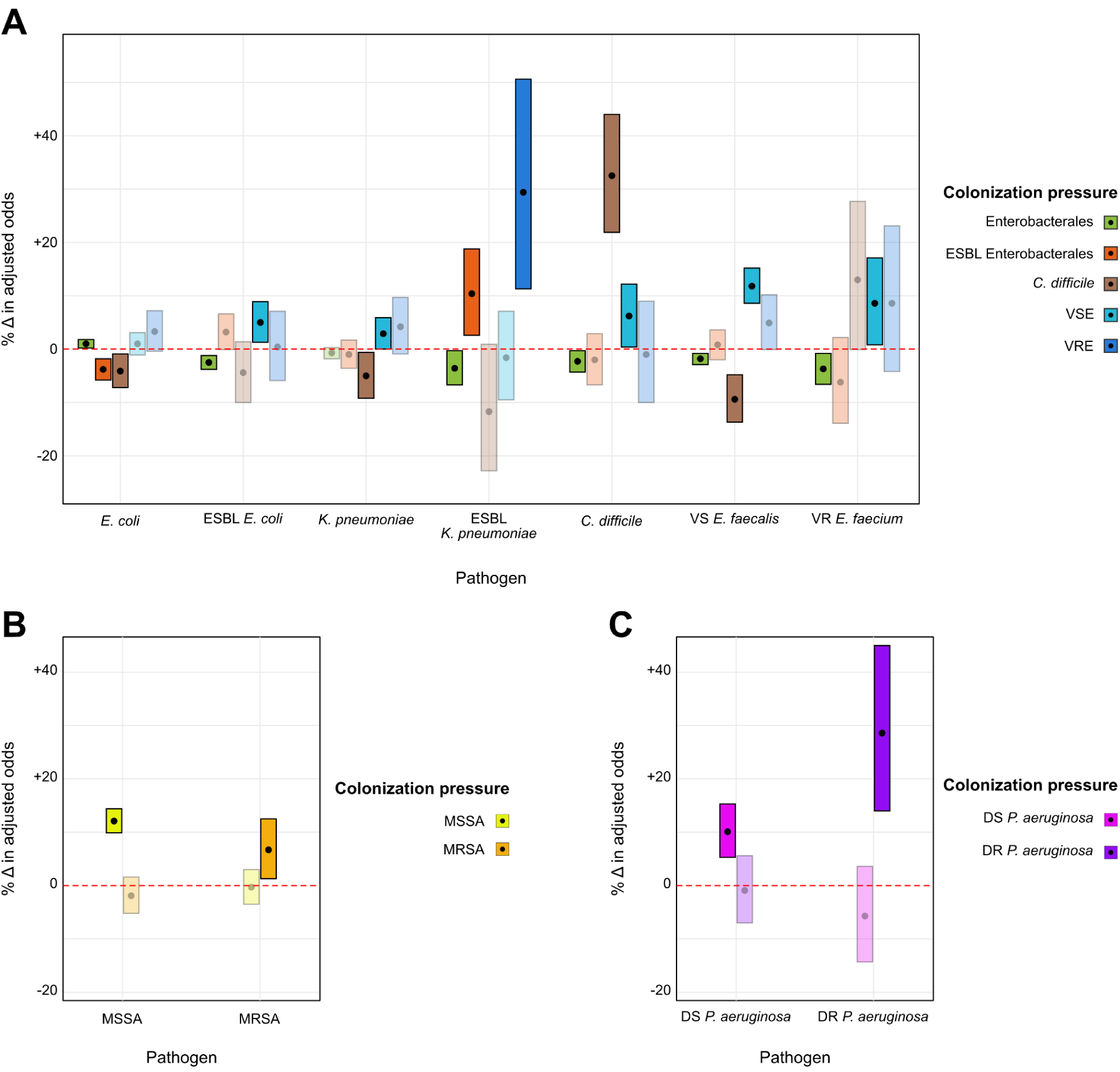
Impact of colonization pressure on adjusted odds of hospital acquisition of cognate organisms. A) Impact of CP from enteric organisms on hospital acquisition of an enteric organism. B) Impact of CP from skin flora on hospital acquisition of skin flora. C) Impact of CP from environmental flora on hospital acquisition of environmental flora. Point estimates for effect size are given by black circles. Bars represent the 95% confidence interval around the point estimate. CP features with 95% CIs that do not cross 0% are considered statistically significant and are bolded. Y-axis scale identical across all panels. Abbreviations, VS, vancomycin susceptible; VR, vancomycin resistant; DS, drug susceptible; DR drug resistant.

CP_Enterobacterales_ was associated with a protective effect for hospital acquisition from 5 enteric organisms, including ESBL *E. coli* (-2.5%, 95% CI -3.8% to -1.2%), ESBL *K. pneumoniae* (-3.6%, 95% CI -6.7% to -0.3%), *C. difficile* (-2.3%, 95% CI -4.3% to -0.3%), vancomycin susceptible *E. faecalis* (-1.8%, 95% CI -2.9% to -0.8%), and vancomycin resistant *E. faecium* (-3.7%, 95% CI -6.6% to -0.8%). Similarly, CP*_C. difficile_* had a protective effect for hospital acquisition of drug susceptible *E. coli* (-4.1%, 95% CI -7.2% to -0.9%), *K. pneumoniae* (-5.0%, 95% CI -9.2% to -0.6%) and vancomycin susceptible *E. faecalis* (-9.4%, 95% CI -13.7% to -4.8%).

Among the skin flora, a unit increase in CP_MSSA_ was associated with a +12.1% (95% CI +9.9% to +14.4%) increase in the odds of nosocomial acquisition of MSSA, and a unit increase in CP_MRSA_ was associated with a +6.7% (95% CI +1.3% to +12.5%) increase in the odds of nosocomial acquisition of MRSA. Among environmental flora, CP_PsA-DS_ was associated with a +10.1% (95% CI +5.3% to +15.3%) increase in the odds of hospital acquired drug susceptible *P. aeruginosa* and similarly, CP_PsA-DR_ was associated with a +28.6% (95% CI +14.0% to +45.0%) increase in the odds of hospital acquired drug resistant *P. aeruginosa*.

### Colonization pressure and nosocomial acquisition among non-cognate pairs

Figure 5 shows the adjusted odds of hospital acquisition of an organism due to non-cognate colonization pressures. Eleven of 13 significant associations were protective in these models. For enteric organisms, a unit increase in CP_MRSA_ was associated with a -5.4% (95% CI -8.3% to -2.2%) and -5.1% (95% CI -8.9% to -1.2%) decrease in the odds of nosocomial acquisition of drug susceptible *E. coli* and *K. pneumoniae*, respectively. A unit increase in CP_MSSA_ was associated with a -7.9% (95% CI -15.1% to -0.2%) decrease in the odds of ESBL *K. pneumoniae*, while a unit increase in CP_PsA-DR_ was associated with a -10.0% (95% CI -15.6% to -4.0%) decrease in the odds of vancomycin susceptible *E. faecalis*.

**Figure 5:**
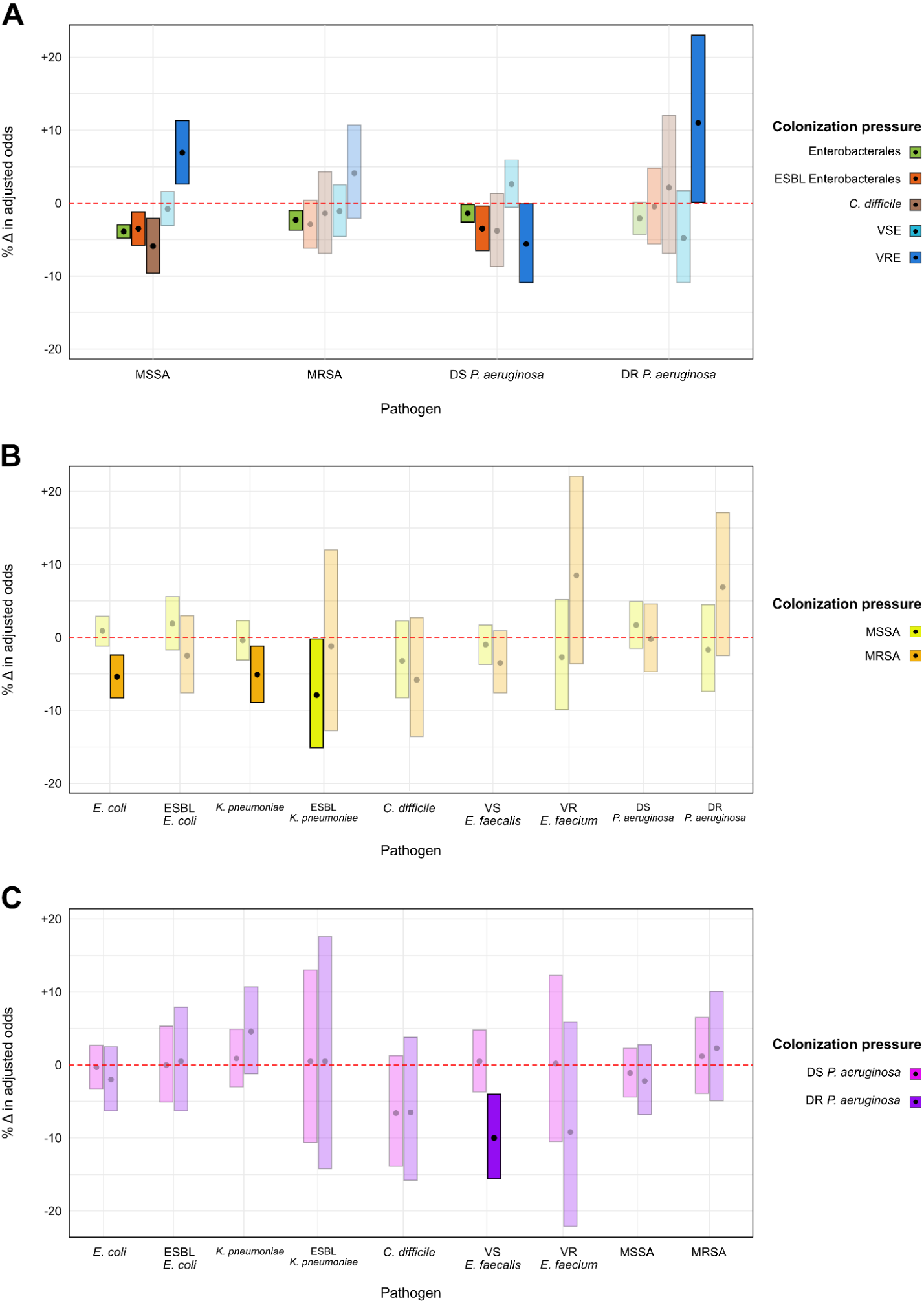
Impact of colonization pressure on adjusted odds of hospital acquisition of non-cognate organisms. A) Impact of CP from enteric organisms on hospital acquisition of skin and environmental organisms. B) Impact of CP from skin flora on hospital acquisition of enteric and environmental organisms. C) Impact of CP from environmental flora on hospital acquisition of enteric and skin organisms. Point estimates for effect size are given by black circles. Bars represent the 95% confidence interval around the point estimate. CP features with 95% CIs that do not cross 0% are considered statistically significant and are bolded. Y-axis scale identical across all plots. Abbreviations, VS, vancomycin susceptible; VR, vancomycin resistant; DS, drug susceptible; DR drug resistant.

Among the skin flora, the odds of hospital acquisition of MSSA were reduced for unit increases in CP_Enterobacterales_ (-3.9%, 95% CI -4.8% to -3.0%), CP_ESBL_ (-3.5%, 95% CI -5.8% to -1.2%), and CP*_C. difficile_* (-5.9%, 95% CI -9.6% to -2.1%), while the odds of MRSA were reduced for unit increases in CP_Enterobacterales_ (-2.3%, 95% CI -3.7% to -1.0%). For the environmental flora, the odds of drug susceptible *P. aeruginosa* decreased with unit increases in CP_Enterobacterales_ (-1.4%, 95% CI -2.6% to -0.2%), CP_ESBL_ (-3.5%, 95% CI -6.5% to -0.4%), and CP_VRE_ (-5.6%, 95% CI -10.9% to -0.1%). Two positive associations were noted, both related to unit increases in CP_VRE_, MSSA (+6.9%, 95% CI +2.6% to +11.3%) and drug resistant *P. aeruginosa* (+11.0%, 95% CI +0.1% to +23.0%).

### Prediction of HAI using colonization pressure

AUROCs for XGB models using CP and Elixhauser index to predict nosocomial acquisition ranged from 0.56 (95% CI 0.49 to 0.62) for ESBL *K. pneumoniae* to 0.63 (95% CI 0.60 to 0.67) for *C. difficile*. Positive predictive values ranged from 33% for MRSA to 50% for *C. difficile*, and negative predictive values ranged from 61% for drug resistant *P. aeruginosa* to 75% for MSSA. Likelihood ratio positive values ranged from 1.05 for MRSA to 1.54 for *C. difficile*, while likelihood ratio negative values ranged from 0.97 for MRSA to 0.68 for *C. difficile*. The full distribution of metrics for the conditional logistic regression and XGBoost prediction models is found in supplementary table S8. Mean SHAP values for conditional logistic regression and XGB models are located in supplementary table S9 and SHAP plots are located in supplementary figures S6 through S11.

## DISCUSSION

Using routinely collected EHR data, we show that a patient’s odds of acquiring a potential pathogen during a hospitalization is associated with the burden of infection among that patient’s ward co-occupants. Associations were noted for drug resistant as well as drug susceptible organisms, between those that typically inhabit the same as well as distinct niches, and included direct as well as indirect relationships. Our findings reinforce the hypothesis that colonization pressure is a proxy of a unit’s microbiome, and open up new and potentially important avenues of research for hospital epidemiology and microbial ecology.

There is a large body of work describing the risks of infection conferred by the built environment and healthcare delivery. Metagenomic investigations into the biogeography of indoor environments and hospital rooms point to bidirectional transmission between room occupants and their room^21–24^, with adverse clinical outcomes of room-to-occupant transmission concentrated in certain vulnerable subpopulations such as neonates^25,26^. Our expanded scope of analysis suggests room-to-occupant transmission may be widespread throughout the hospital. Health care epidemiology analyses provide a complementary perspective by defining the role of healthcare workers as vectors for transmission^3,24,27^ and quantifying individual level risk factors for hospital acquired infection^28^. While both fields have yielded critical insights, a common weakness is the use of variables that cannot be easily operationalized for real time surveillance. A smaller number of studies have examined colonization pressure, which links microbial ecology with epidemiology, but they are limited to cognate relationships and focus almost exclusively on intensive care units^10,12,29–31^ and high-risk clones that are well adapted for survival and transmission in hospital wards^10,32^. Our study reaffirms the findings from these bodies of work, while highlighting novel relationships for drug susceptible organisms and potentially inverse interactions using multivariate models.

High-risk clones are thought to be successful due to the accumulation of phenotypic traits that confer a survival advantage under the selective pressure of the hospital environment. These elements include resistance to antimicrobials and disinfectants, the ability to form biofilms and the expression of virulence factors that permit survival under pressure from microbial competitors and the host immune system^33^. Our findings suggest that while these factors may lead clones to transmit more efficiently than strains that lack them, they may not be necessary for hospital acquisition, particularly when the burden of colonization within a unit is high. It is notable that despite having weaker associations, the absolute burden attributable to hospital acquired drug susceptible organisms was several times greater than that due to drug resistant organisms in our study. This may be the result of a lack of targeted infection control measures for this organism group. In our study, this is reflected by the higher mean colonization pressures for drug susceptible organisms (Enterobacterales, VSE, MSSA, drug susceptible *P. aeruginosa*) than for drug resistant organisms (ESBLs, VRE, MRSA and drug resistant *P. aeruginosa*).

Nosocomial acquisition for several organisms had an inverse relationship with colonization pressure, meaning model-adjusted CP values were higher in controls than in cases. This was apparent for both cognate and non-cognate pairs, though it occurred more often in the latter. There are several potential explanations for this. The first two involve confounding between comparators due to differences in ward characteristics or patient characteristics. For instance, if cases were on wards with fewer double occupancy rooms^34^, or higher rates of contact precautions, then rates of nosocomial acquisition would be expected to be lower. A similar outcome could be seen if a higher proportion of controls had unmeasured patient-specific features associated with nosocomial acquisition, such as a specific type of surgery, hardware or intubation. Though only true randomization would eliminate bias entirely from the analysis, both of these scenarios are somewhat unlikely as we applied stringent matching criteria, including across 14 classes of antibiotics. This serves two purposes. First, it reduces differences in selective pressures that would promote silent carriage, and second, because single and combination antibiotic therapies have a finite number of indications, fine-grained matching effectively clusters similar types of patients together onto similar wards. This is supported by the similar distributions of individual Elixhauser categories between cases and controls, despite not being included in the match.

A third possible explanation for the inverse relationship is that contamination of a ward by one organism may prevent other organisms from establishing themselves. This may be due to repeated contamination by colonized hosts on a given floor, or alternatively, intra- and inter-species competition on hospital environmental surfaces. The concept of colonization resistance and competition has been described *in vitro* and for niches across the human body^35–38^. Further research is necessary to prove whether founder effects and other competitive mechanisms take place on high touch surfaces, sinks and bathrooms within hospital units. A causal relationship offers the possibility of leveraging competition between organisms to reduce rates of transmission and hospital-acquired infection from difficult to treat pathogens^39^. On a more pragmatic level, it increases the utility of passive surveillance of unit-level CP.

In addition to supporting reproducibility, our goal with releasing an extensively annotated analytic pipeline and de-identified patient-level data is to allow other hospital systems to adapt and improve on our methods. Our hope is for the research community to collectively build a generalizable, automated, real-time risk assessment tool for nosocomial pathogen acquisition that functions across the hospital, for any range of organisms, and eventually across different health systems. Such a tool could be used to study colonization pressure on a broader scale, which we hope will generate an evidence base for pre-emptive measures to prevent infections or enact closer surveillance on wards that cross a pre-designated threshold. It is important to note however, that despite the advantages of using a machine learning approach to maximize predictive power, our XGBoost models had low positive and negative predictive value for hospital acquisition. This may be explained by small sample sizes and the fact that colonization pressure is one of many factors that must be included in real-time models to support decision-making.

The other major strength of this study is our incorporation of a wide range of common organisms into our CP models. This allows us to estimate the odds of hospital acquisition for a given organism while accounting for the complex ecology of a hospital unit, and to identify interactions that may be occurring across species and niches. To our knowledge this is the first study to utilize this strategy. Future work aims to improve our model’s predictive performance by addition of host-specific factors and integration with clinical pathogen whole genome sequencing data to better define transmission events. Model performance will be improved by including data from other hospital systems, which will bolster sample size and generalizability.

Our study has a number of limitations. Our modest sample sizes restrict the number of features that could be included in the models. This reflects a decision to apply very strict inclusion criteria for cases and controls, thereby providing the cleanest possible comparison for estimating the impact of hospital environment on nosocomial acquisition using observational data. For instance, the use of highly granular matching of antibiotic exposures by both class and burden of exposure helps ensure that selective pressures from systemic antimicrobials are closely aligned for cases and controls. Furthermore, restricting the observation period to the first room prevents confounding that may occur if the patient was exposed to another hospital unit during the same hospitalization. Nevertheless, there is the possibility of residual confounding. The results represent a proof-of-principle that CP impacts nosocomial acquisition, but we do not quantify its importance relative to other factors. Another limitation is our inability to identify silent carriage of a target organism, which could confound the relationship between CP and hospital acquisition. This stems partly from the fact that we have a lack of insight into events (microbiology specimens, antibiotic prescriptions) that occur outside of our healthcare system. We have reduced this possibility by excluding patients with evidence of the target organism in the previous 6 or 12 months. Finally, our data come from a single healthcare system in the northeast US and caution is warranted when extrapolating to other geographic areas and healthcare systems that may differ in terms of patient demographics, antibiotic consumption and infection control practices. This could be surmounted by combining data across multiple health systems using a common data model or a federated learning approach.

In conclusion, our data affirm the importance of colonization pressure at the hospital unit level as a risk factor for acquisition of common pathogens. We quantify the effect for 11 drug susceptible and drug resistant organisms, inhabiting 3 distinct niches, and identified direct and inverse relationships with 9 CP organism sets. While more easily treated with antibiotics, the absolute burden of hospital acquired drug susceptible organisms in our health system was four times higher than that for the organisms typically surveilled by infection control programs. Further research is necessary to understand the costs and benefits of applying real-time surveillance of colonization pressure to prevent hospital acquired infection.

## Data Availability

All data produced are available online at Physionet

## SUPPLEMENTARY MATERIAL

**Figure S1:**
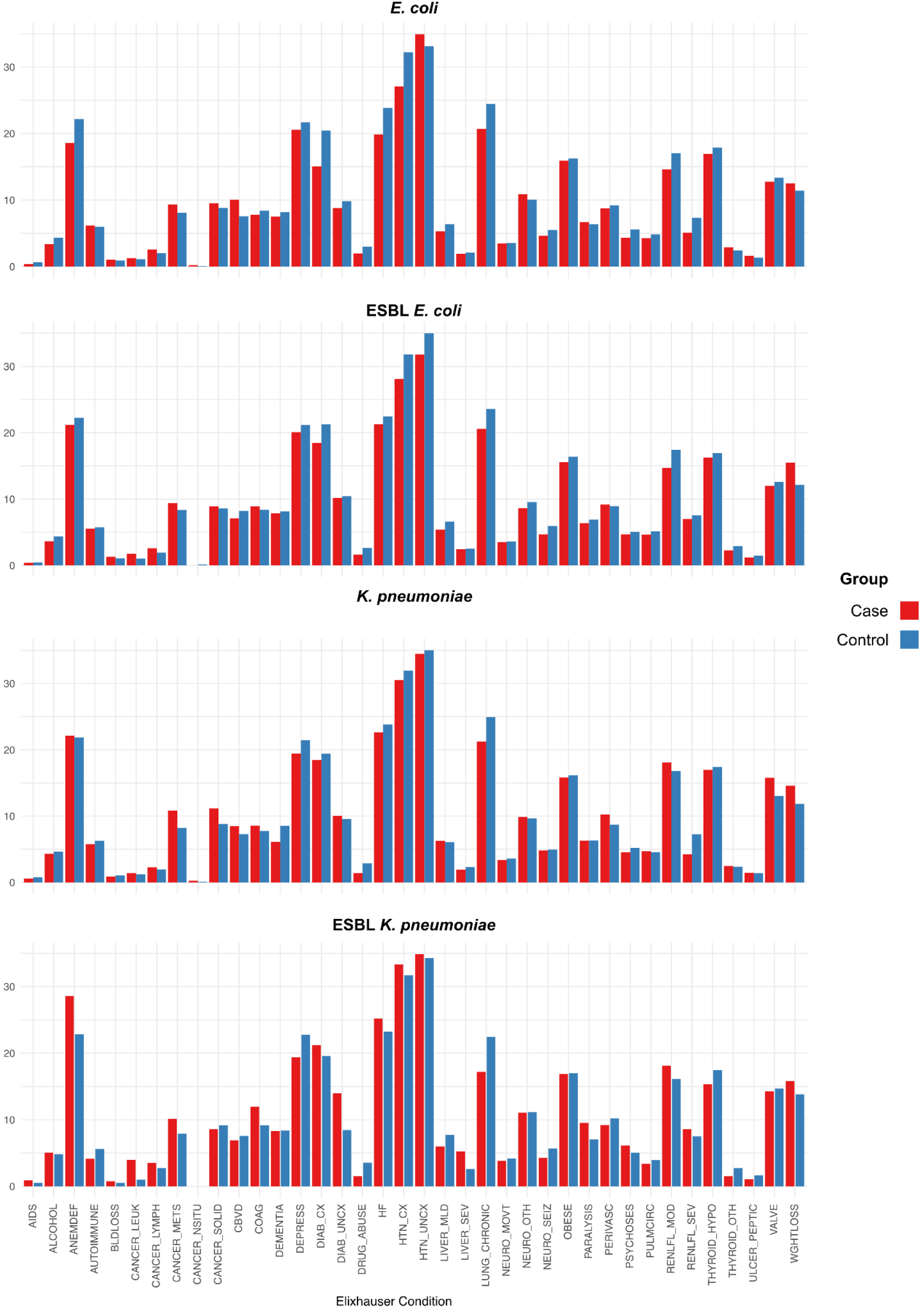
Distribution of individual elixhauser categories for drug susceptible and ESBL *E. coli* and *K. pneumoniae*.

**Figure S2:**
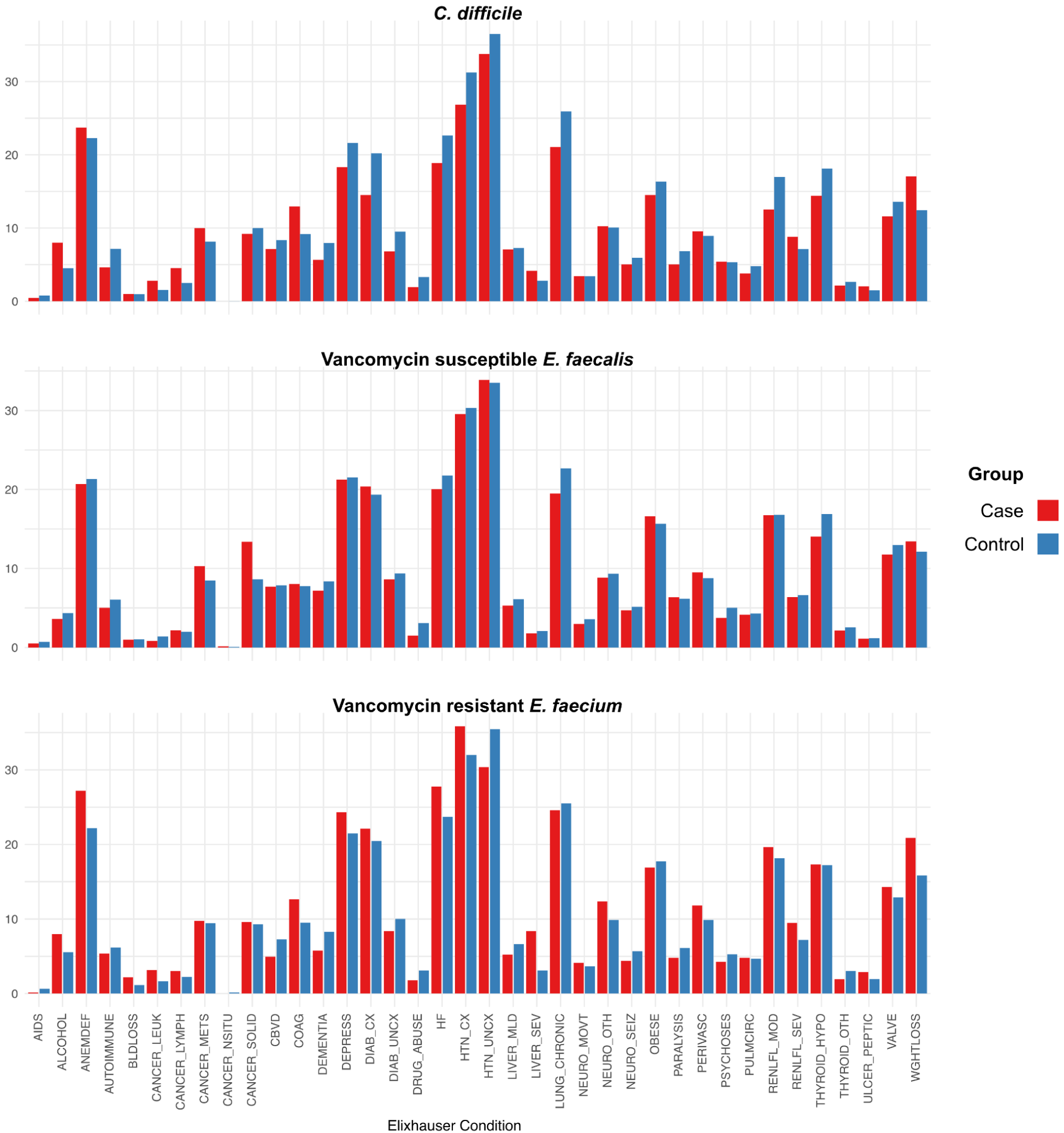
Distribution of individual elixhauser categories for *C. difficile,* vancomycin susceptible *E. faecalis*, and vancomycin resistant *E. faecium*.

**Figure S3:**
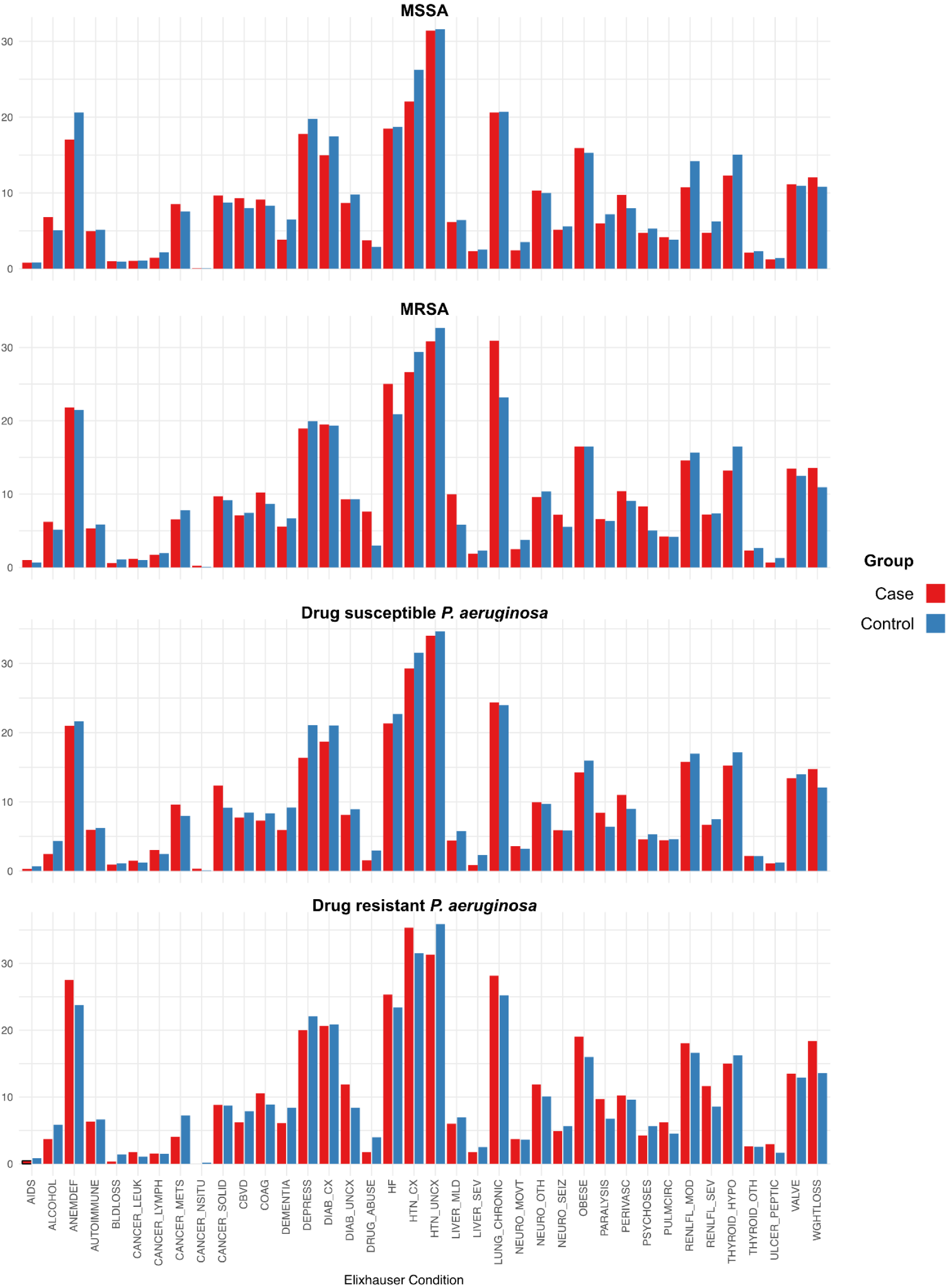
Distribution of individual elixhauser categories for MSSA, MRSA, drug susceptible and drug resistant *P. aeruginosa*.

**Figure S4:**
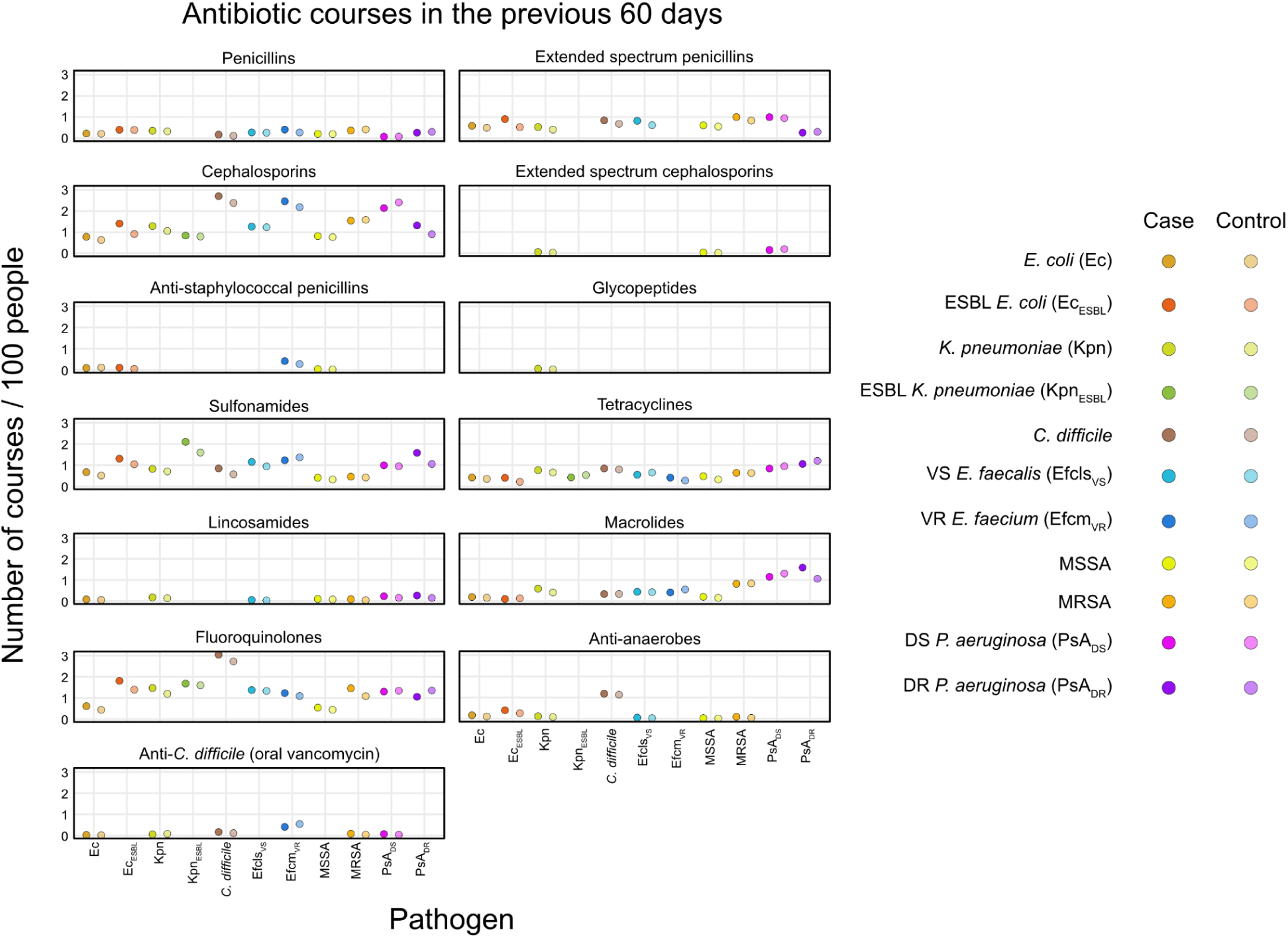
Number of antibiotic courses per 100 people in the previous 60 days by class.

**Figure S5:**
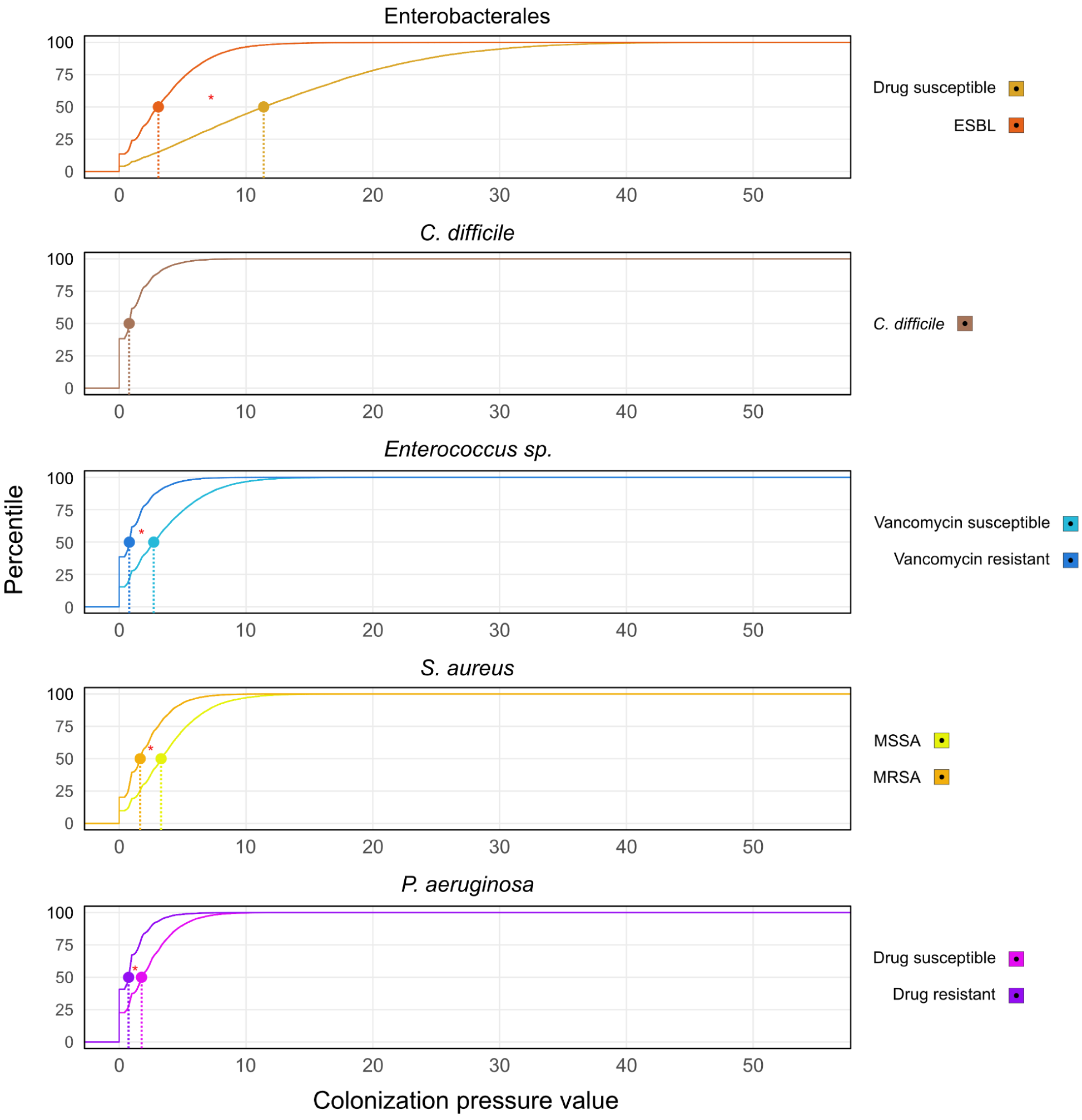
Cumulative distributions of colonization pressure. Vertical line indicates the median value and the red asterisk indicates p < 0.05 for comparison of distributions.

**Figure S6:**
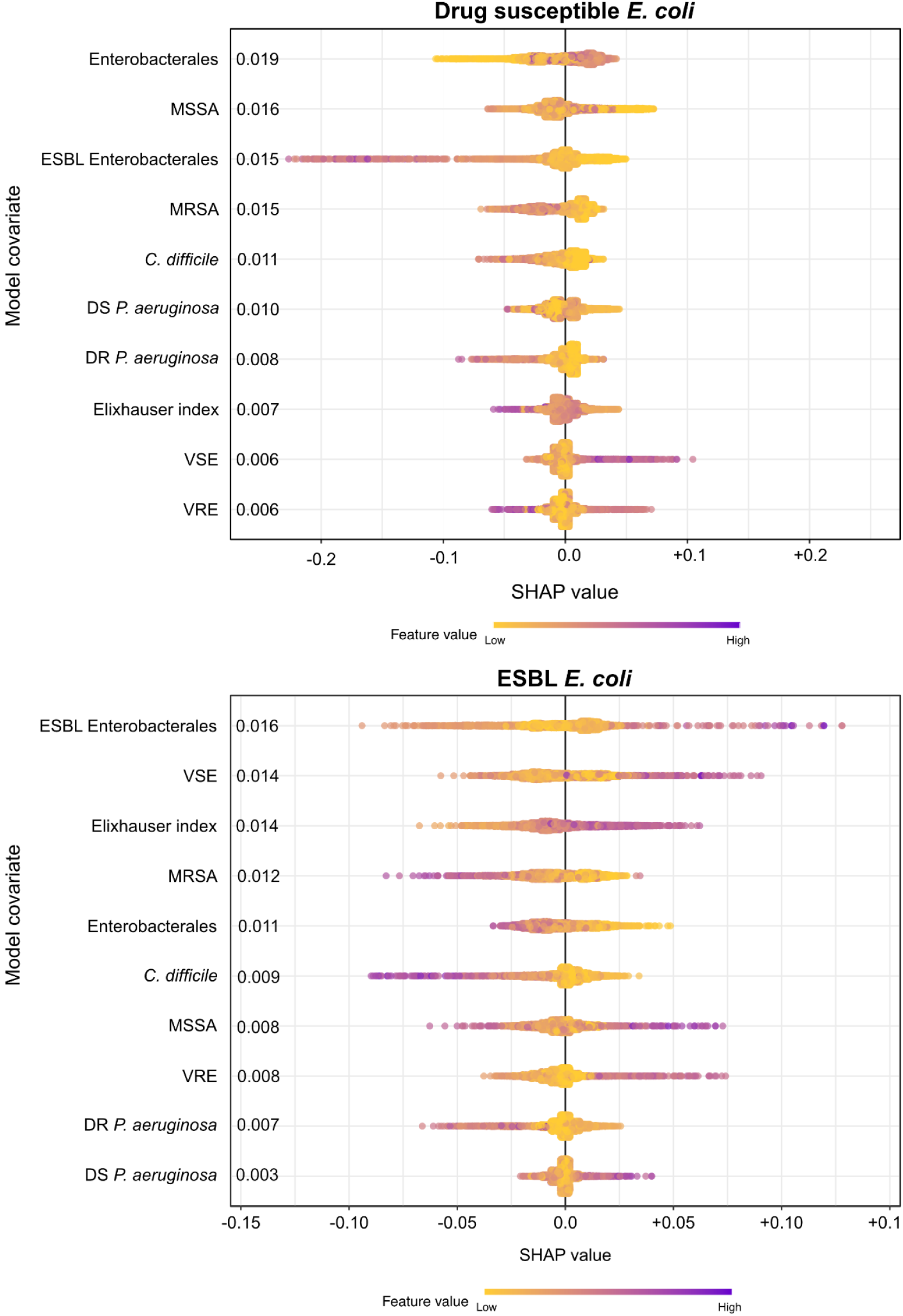
SHAP values for XGB models predicting nosocomial acquisition of A) drug susceptible and B) ESBL *E. coli*. Mean SHAP value on left of plot. All features represent organism-specific colonization pressure except for Elixhauser index. Features are ranked by importance.

**Figure S7:**
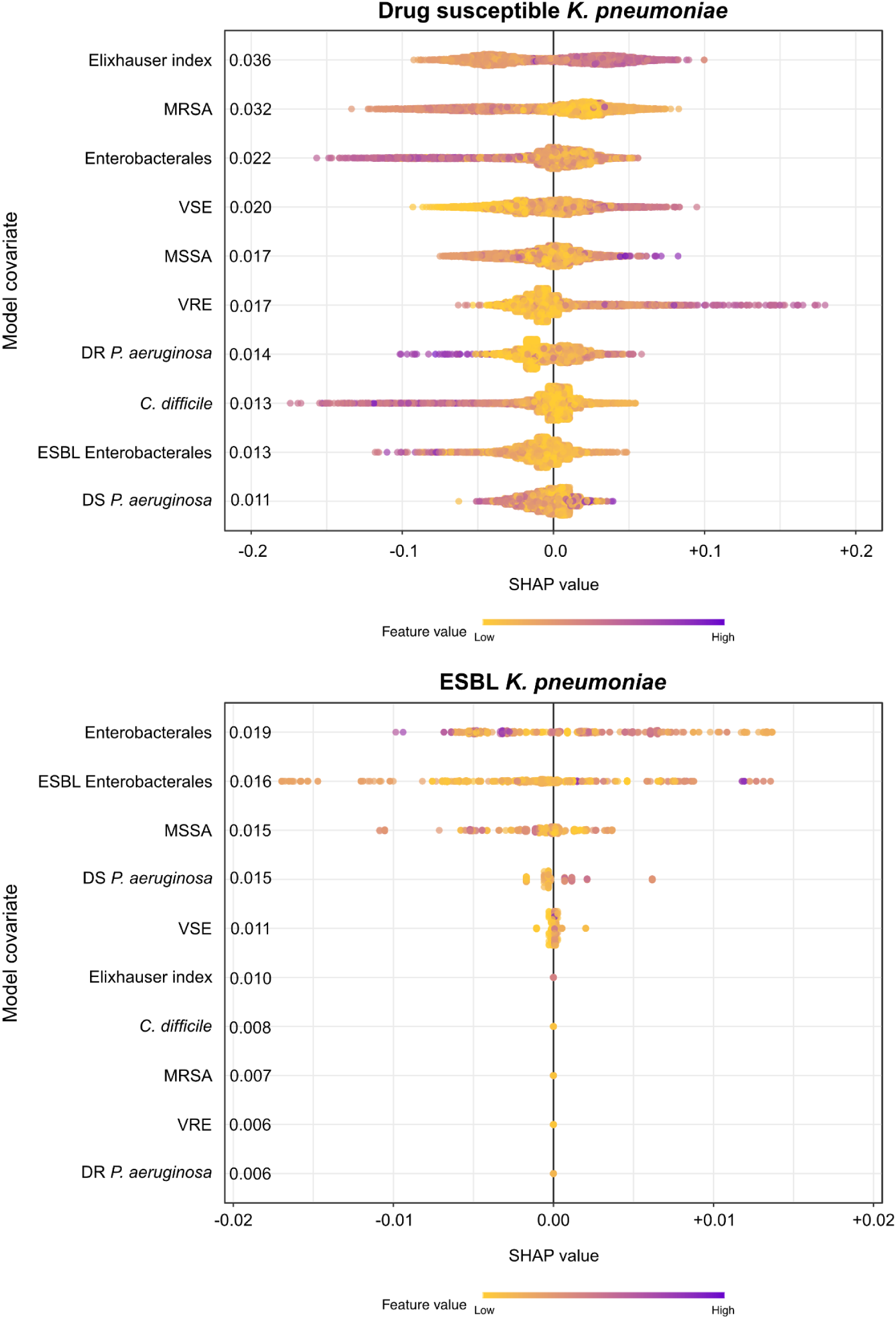
SHAP values for XGB models predicting nosocomial acquisition of A) drug susceptible and B) ESBL *K.pneumoniae*. Mean SHAP value on left of plot. All features represent organism-specific colonization pressure except for Elixhauser index. Features are ranked by importance.

**Figure S8:**
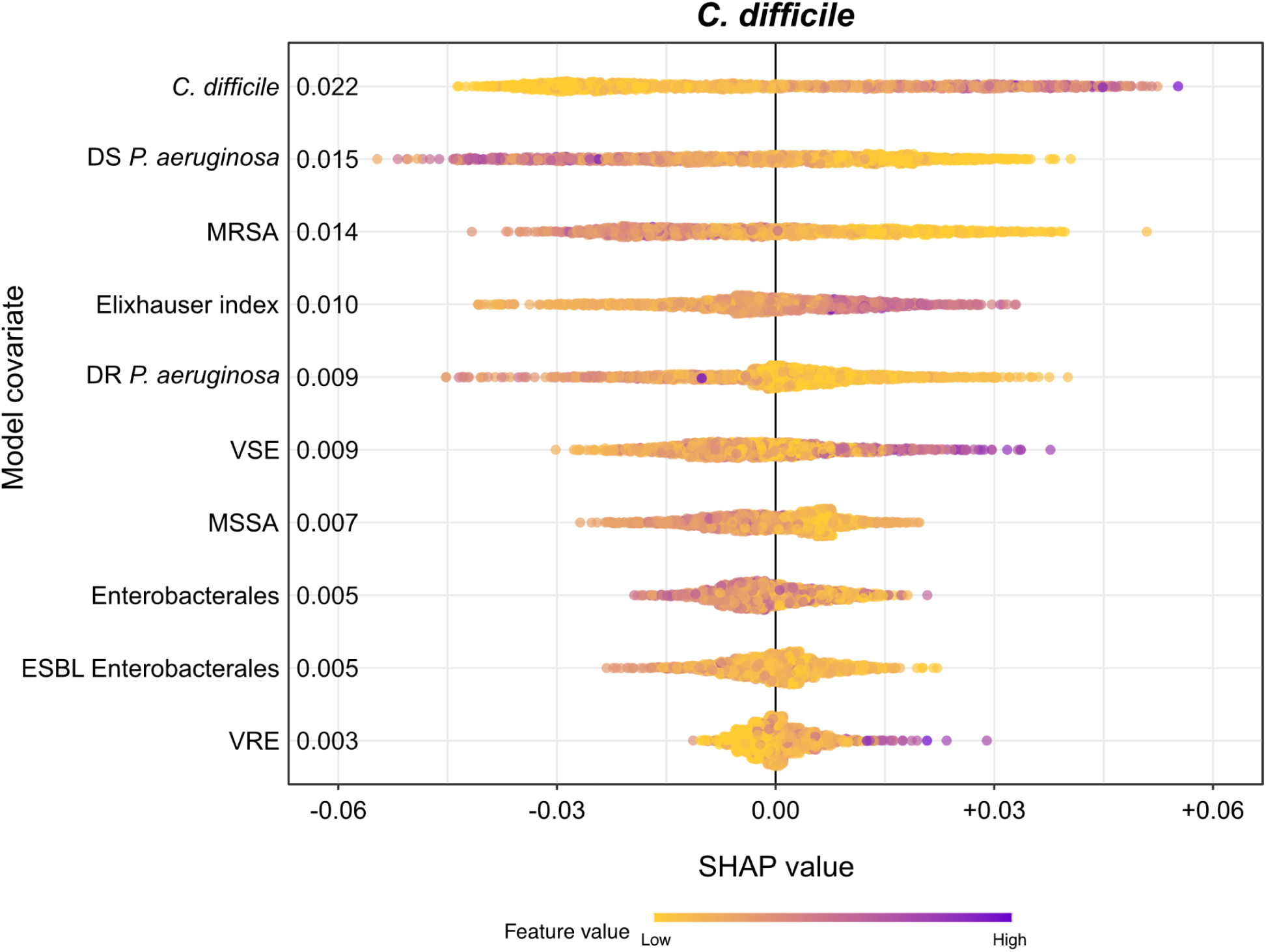
SHAP values for XGB models predicting nosocomial acquisition of *C. difficile*. Mean SHAP value on left of plot. All features represent organism-specific colonization pressure except for Elixhauser index. Features are ranked by importance.

**Figure S9:**
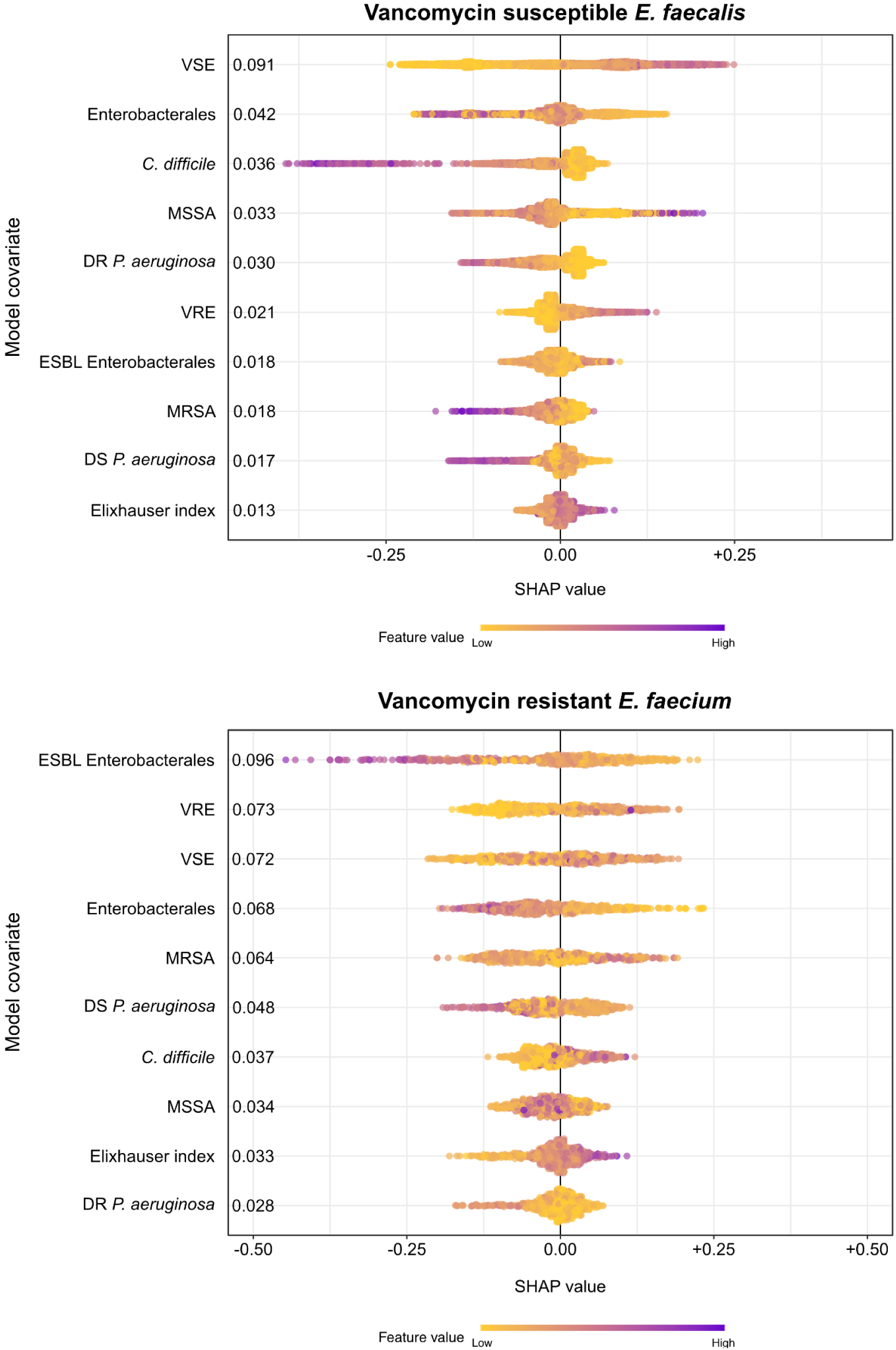
SHAP values for A) vancomycin susceptible *E. faecalis* and B) vancomycin resistant *E. faecium*. Mean SHAP value on left of plot. All features represent organism-specific colonization pressure except for Elixhauser index. Features are ranked by importance.

**Figure S10:**
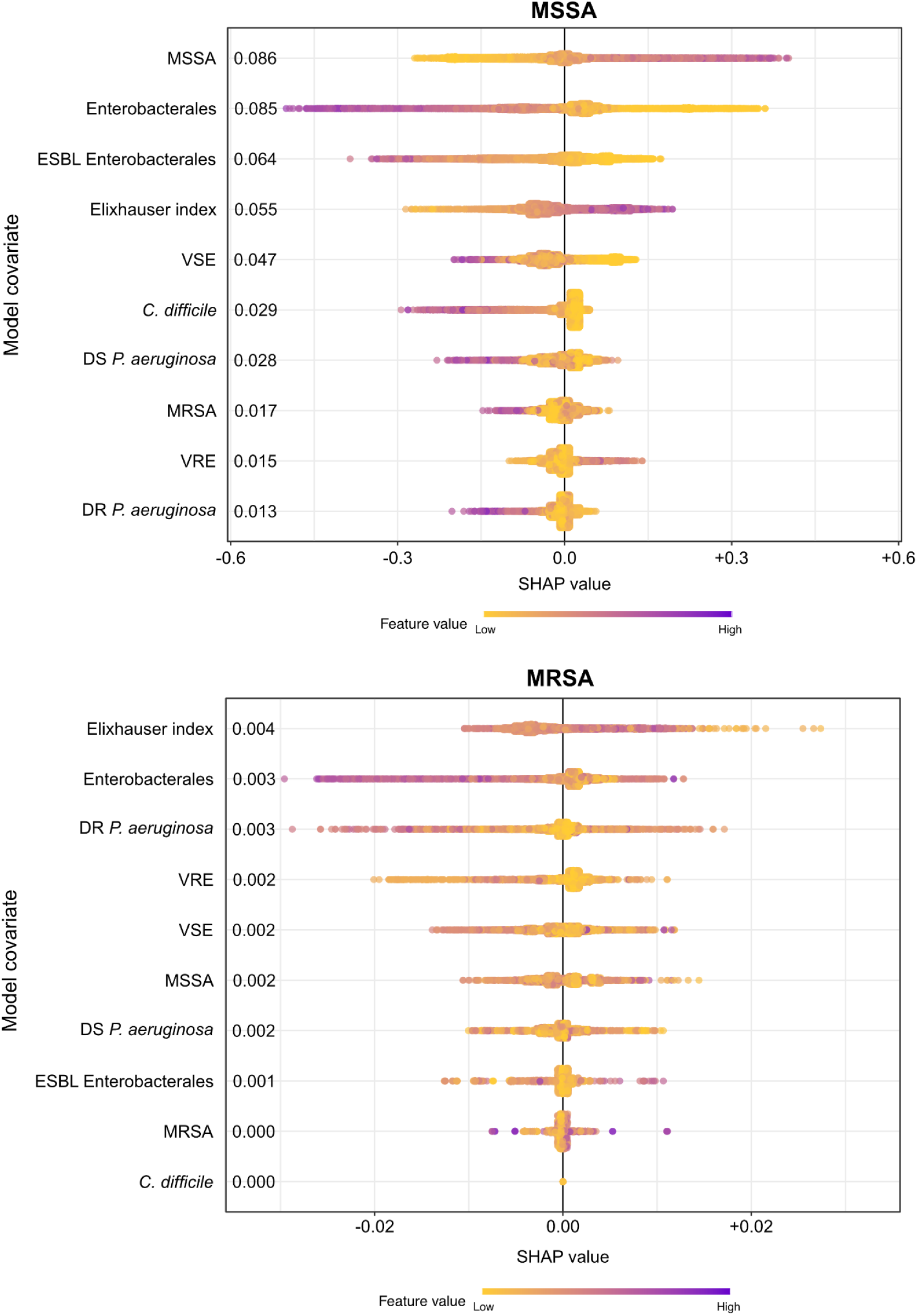
SHAP values for XGB models predicting nosocomial acquisition of A) MSSA and B) MRSA. Mean SHAP value on left of plot. All features represent organism-specific colonization pressure except for Elixhauser index. Features are ranked by importance.

**Figure S11:**
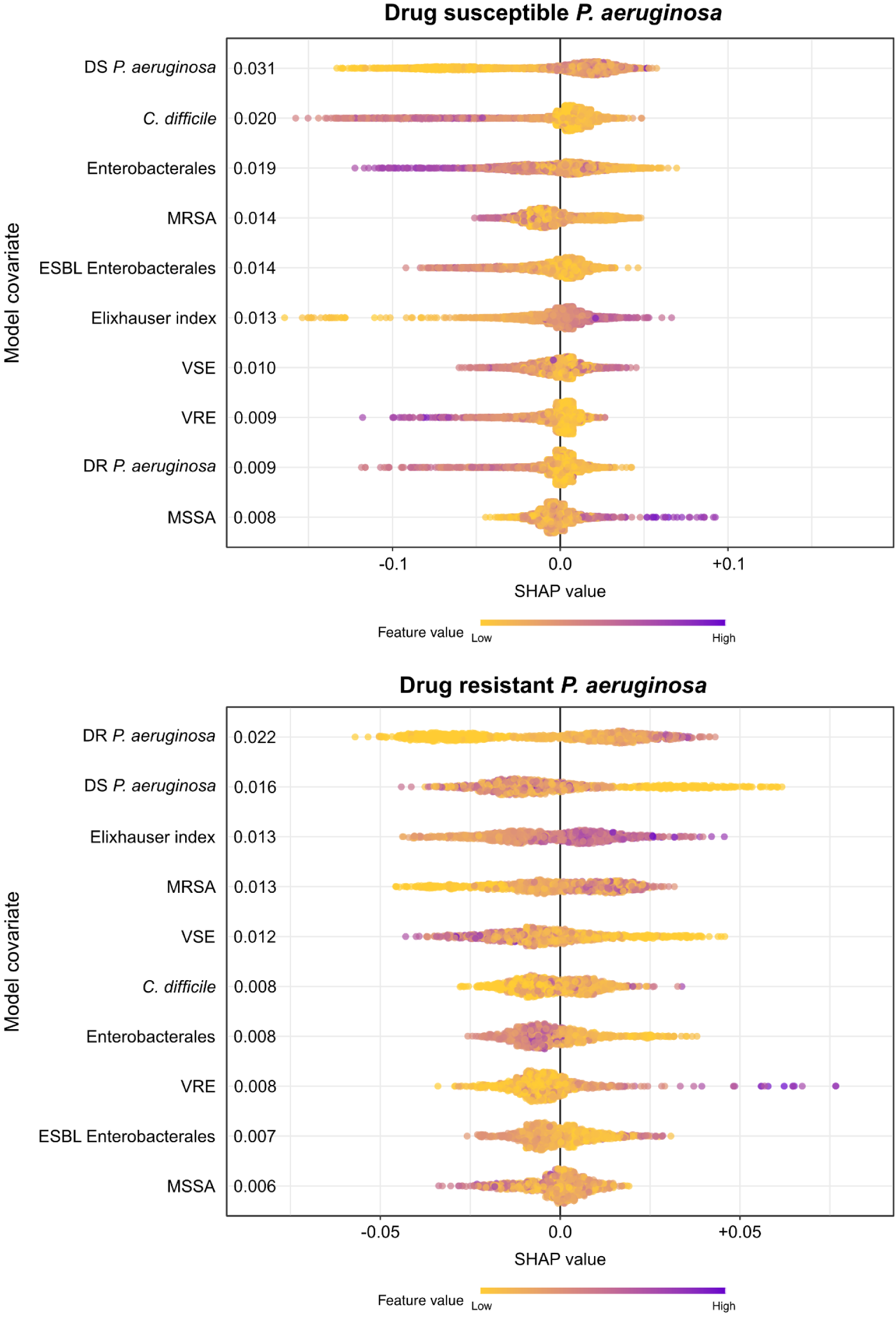
SHAP values for XGB models predicting nosocomial acquisition of A) drug susceptible and B) drug resistant *P. aeruginosa*. Mean SHAP value on left of plot. All features represent organism-specific colonization pressure except for Elixhauser index. Features are ranked by importance.

**Table S1:**
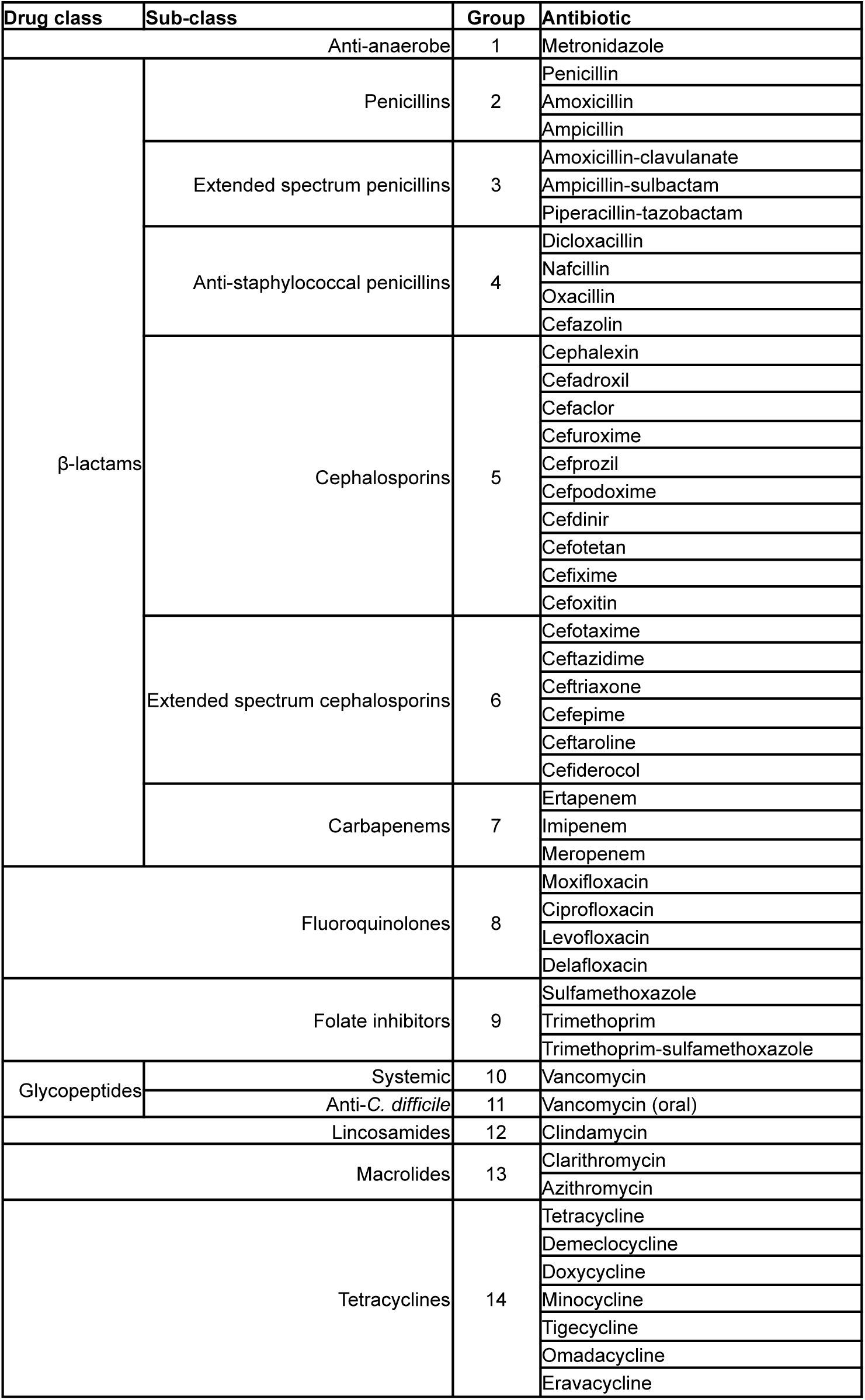
Antibiotic categories.

**Table S2:** Flow diagrams for all cohort cases. Abbreviations, VS, vancomycin susceptible; VR, vancomycin resistant; DS, drug susceptible; DR, drug resistant

| Pathogen | Total admitted patients with pathogen of interest | No antibiotic treatment between -7 days to +3 days | No prior encounters within 90 days | Pathogens of interest identified after day 3 and before day 30 | Drop cases not in the first room | Drop cases with prior infection | Controls matched to cases |
| --- | --- | --- | --- | --- | --- | --- | --- |
| *E. coli* | 64,492 | 44,653 | 30,344 | 7,594 | 6,137 | 5,701 | 3,585 |
| ESBL *E. coli* | 16,482 | 10,462 | 5,890 | 1,783 | 1,323 | 1,203 | 996 |
| *K. pneumoniae* | 26,848 | 17,627 | 10,245 | 2,789 | 2,165 | 2,037 | 1,704 |
| ESBL *K. pneumoniae* | 6,662 | 4,033 | 1,813 | 639 | 437 | 404 | 238 |
| VS *E. faecalis* | 34,875 | 22,763 | 13,209 | 4,030 | 3,031 | 2,864 | 1,823 |
| VR *E. faecium* | 8,461 | 2,119 | 2,054 | 837 | 521 | 482 | 244 |
| *C. difficile* | 13,053 | 8,659 | 4,342 | 1,883 | 1,415 | 1,365 | 592 |
| MSSA | 41,275 | 26,441 | 16,972 | 4,593 | 3,573 | 3,369 | 2,952 |
| MRSA | 21,503 | 12,843 | 7,225 | 1,897 | 1,443 | 1,305 | 1,101 |
| DS *P. aeruginosa* | 21,257 | 12,664 | 7,115 | 2,427 | 1,812 | 1,694 | 1,309 |
| DR *P. aeruginosa* | 9,312 | 5,373 | 2,729 | 832 | 627 | 561 | 379 |

**Table S3:** Baseline characteristics for *E. coli*, ESBL *E. coli* cohorts. Abbreviations, SE, standard error; IQR, Interquartile Range.

| Feature | *E. coli* |  | ESBL *E. coli* |  |
| --- | --- | --- | --- | --- |
|  | Case | Control | Case | Control |
| Sample size | 3,585 | 5,693 | 996 | 2,299 |
| Demographics |  |  |  |  |
| Age (mean, SE) | 66.5 +/- 0.3 | 66.7 +/- 0.3 | 66.7 +/- 0.6 | 68.3 +/- 0.4 |
| Gender (% female) | 71.1 | 69.6 | 61.7 | 63.0 |
| Elixhauser index (mean, SE) | 9.7 +/- 0.3 | 9.3 +/- 0.2 | 10.8 +/- 0.5 | 9.6 +/- 0.3 |
| Prior surgery (%) | 13.0 | 10.8 | 15.9 | 13.0 |
| Length of Stay (median, IQR) | 4.5 (2.9, 7.9) | 3.8 (2.8, 5.0) | 5.1 (3.5, 8.5) | 3.9 (2.9, 5.7) |
| Matching Duration (median, IQR) | 3.9 (2.9, 5.6) | 3.8 (2.8, 5.0) | 4.2 (3.2, 6.1) | 3.9 (2.9, 5.7) |
| Time to Infection (median, IQR) | 9.0 (4.9, 17.2) | 0.0 (0.0, 0.0) | 8.8 (5.1, 17.6) | 0.0 (0.0, 0.0) |
| Prior antimicrobials (%) |  |  |  |  |
| β-lactams |  |  |  |  |
| Penicillins | 0.2 | 0.2 | 0.4 | 0.4 |
| Extended spectrum penicillins | 0.6 | 0.5 | 0.9 | 0.5 |
| Cephalosporins | 0.8 | 0.6 | 1.4 | 0.9 |
| Extended spectrum cephalosporins | 0.0 | 0.0 | 0.0 | 0.0 |
| Anti-Staphylococcal β-lactams | 0.1 | 0.1 | 0.1 | 0.0 |
| Other cell wall active agents |  |  |  |  |
| Glycopeptides | 0.0 | 0.0 | 0.0 | 0.0 |
| DNA synthesis inhibitors |  |  |  |  |
| Sulfonamides | 0.7 | 0.5 | 1.3 | 1.0 |
| Protein synthesis inhibitors |  |  |  |  |
| Fluoroquinolones | 0.6 | 0.4 | 1.8 | 1.4 |
| Tetracyclines | 0.4 | 0.4 | 0.4 | 0.2 |
| Macrolides | 0.2 | 0.2 | 0.1 | 0.1 |
| Lincosamides | 0.1 | 0.1 | 0.0 | 0.0 |
| Anti-anaerobic antibiotics |  |  |  |  |
| Metronidazole | 0.2 | 0.1 | 0.4 | 0.3 |
| Oral vancomycin | 0.0 | 0.0 | 0.0 | 0.0 |

**Table S4:** Baseline characteristics for *K. pneumoniae*, and ESBL *K. pneumoniae* cohorts. Abbreviations, SE, standard error; IQR, Interquartile Range.

| Feature | *K. pneumoniae* |  | ESBL *K. pneumoniae* |  |
| --- | --- | --- | --- | --- |
|  | Case | Control | Case | Control |
| Sample size | 1,704 | 3,684 | 238 | 376 |
| Demographics |  |  |  |  |
| Age (mean, SE) | 67.8 +/- 0.4 | 68.4 +/- 0.3 | 69.3 +/- 1.0 | 69.0 +/- 0.9 |
| Gender (% female) | 58.2 | 60.4 | 50.4 | 53.5 |
| Elixhauser index (mean, SE) | 11.6 +/- 0.4 | 9.4 +/- 0.3 | 12.9 +/- 1.0 | 10.2 +/- 0.8 |
| Prior surgery (%) | 14.4 | 13.6 | 8.8 | 8.1 |
| Length of Stay (median, IQR) | 5.3 (3.5, 8.9) | 4.0 (3.0, 5.6) | 6.3 (3.7, 12.1) | 4.8 (3.2, 7.9) |
| Matching Duration (median, IQR) | 4.5 (3.2, 6.0) | 4.0 (3.0, 5.6) | 4.9 (3.5, 8.0) | 4.8 (3.2, 7.9) |
| Time to Infection (median, IQR) | 8.8 (5.2, 17.1) | 0.0 (0.0, 0.0) | 11.4 (6.1, 19.0) | 0.0 (0.0, 0.0) |
| Prior antimicrobials (%) |  |  |  |  |
| β-lactams |  |  |  |  |
| Penicillins | 0.4 | 0.3 | 0.0 | 0.0 |
| Extended spectrum penicillins | 0.5 | 0.4 | 0.0 | 0.0 |
| Cephalosporins | 1.3 | 1.1 | 0.8 | 0.8 |
| Extended spectrum cephalosporins | 0.1 | 0.0 | 0.0 | 0.0 |
| Anti-Staphylococcal β-lactams | 0.0 | 0.0 | 0.0 | 0.0 |
| Other cell wall active agents |  |  |  |  |
| Glycopeptides | 0.1 | 0.0 | 0.0 | 0.0 |
| DNA synthesis inhibitors |  |  |  |  |
| Sulfonamides | 0.8 | 0.7 | 2.1 | 1.6 |
| Protein synthesis inhibitors |  |  |  |  |
| Fluoroquinolones | 1.5 | 1.2 | 1.7 | 1.6 |
| Tetracyclines | 0.8 | 0.7 | 0.4 | 0.5 |
| Macrolides | 0.6 | 0.4 | 0.0 | 0.0 |
| Lincosamides | 0.2 | 0.1 | 0.0 | 0.0 |
| Anti-anaerobic antibiotics |  |  |  |  |
| Metronidazole | 0.1 | 0.1 | 0.0 | 0.0 |
| Oral vancomycin | 0.1 | 0.1 | 0.0 | 0.0 |

**Table S5:** Baseline characteristics for vancomycin susceptible *E. faecalis*, vancomycin resistant *E. faecium*, and *C. difficile* cohorts. Abbreviations, SE, standard error; IQR, Interquartile Range.

| Feature | Vancomycin susceptible<br>*E. faecalis* |  | Vancomycin resistant<br>*E. faecium* |  | *C. difficile* |  |
| --- | --- | --- | --- | --- | --- | --- |
|  | Case | Control | Case | Control | Case | Control |
| Sample size | 1,823 | 3,082 | 244 | 366 | 592 | 881 |
| Demographics |  |  |  |  |  |  |
| Age (mean, SE) | 65.7 +/- 0.5 | 65.8 +/- 0.4 | 70.7 +/- 0.9 | 70.7 +/- 0.8 | 68.9 +/- 0.6 | 70.7 +/- 0.5 |
| Gender (% female) | 53.3 | 52.8 | 59.0 | 60.9 | 56.4 | 58.4 |
| Elixhauser index (mean, SE) | 10.3 +/- 0.4 | 9.6 +/- 0.3 | 14.3 +/- 1.0 | 12.0 +/- 0.8 | 13.4 +/- 0.7 | 9.9 +/- 0.5 |
| Prior surgery (%) | 17.9 | 17.7 | 13.9 | 11.7 | 14.7 | 12.9 |
| Length of Stay (median, IQR) | 4.6 (3.0, 7.9) | 3.8 (2.8, 5.0) | 7.3 (4.5, 13.7) | 4.8 (3.8, 7.6) | 5.8 (3.2, 11.0) | 4.0 (2.9, 6.0) |
| Matching Duration (median, IQR) | 4.0 (3.0, 5.8) | 3.8 (2.8, 5.0) | 5.7 (3.9, 8.9) | 4.8 (3.8, 7.6) | 4.1 (3.1, 7.0) | 4.0 (2.9, 6.0) |
| Time to Infection (median, IQR) | 9.9 (5.4, 18.0) | 0.0 (0.0, 0.0) | 10.5 (6.0, 17.6) | 0.0 (0.0, 0.0) | 9.0 (5.0, 16.8) | 0.0 (0.0, 0.0) |
| Prior antimicrobials (%) |  |  |  |  |  |  |
| β-lactams |  |  |  |  |  |  |
| Penicillins | 0.3 | 0.3 | 0.4 | 0.3 | 0.2 | 0.1 |
| Extended spectrum penicillins | 0.8 | 0.6 | 0.0 | 0.0 | 0.8 | 0.7 |
| Cephalosporins | 1.3 | 1.2 | 2.5 | 2.2 | 2.7 | 2.4 |
| Extended spectrum cephalosporins | 0.0 | 0.0 | 0.0 | 0.0 | 0.0 | 0.0 |
| Anti-Staphylococcal β-lactams | 0.0 | 0.0 | 0.4 | 0.3 | 0.0 | 0.0 |
| Other cell wall active agents |  |  |  |  |  |  |
| Glycopeptides | 0.0 | 0.0 | 0.0 | 0.0 | 0.0 | 0.0 |
| DNA synthesis inhibitors |  |  |  |  |  |  |
| Sulfonamides | 1.2 | 0.9 | 1.2 | 1.4 | 0.8 | 0.6 |
| Protein synthesis inhibitors |  |  |  |  |  |  |
| Fluoroquinolones | 1.4 | 1.3 | 1.2 | 1.1 | 3.0 | 2.7 |
| Tetracyclines | 0.5 | 0.6 | 0.4 | 0.3 | 0.8 | 0.8 |
| Macrolides | 0.4 | 0.4 | 0.4 | 0.5 | 0.3 | 0.3 |
| Lincosamides | 0.1 | 0.0 | 0.0 | 0.0 | 0.0 | 0.0 |
| Anti-anaerobic antibiotics |  |  |  |  |  |  |
| Metronidazole | 0.1 | 0.0 | 0.0 | 0.0 | 1.2 | 1.1 |
| Oral vancomycin | 0.0 | 0.0 | 0.4 | 0.5 | 0.2 | 0.1 |

**Table S6:** Baseline characteristics for MSSA and MRSA cohorts. Abbreviations, SE, standard error; IQR, Interquartile Range.

| Feature | MSSA |  | MRSA |  |
| --- | --- | --- | --- | --- |
|  | Case | Control | Case | Control |
| Sample size | 2,952 | 6,504 | 1,101 | 2,397 |
| Demographics |  |  |  |  |
| Age (mean, SE) | 52.1 +/- 0.5 | 54.7 +/- 0.3 | 58.9 +/- 0.7 | 57.7 +/- 0.5 |
| Gender (% female) | 44.6 | 47.4 | 44.0 | 45.5 |
| Elixhauser index (mean, SE) | 8.8 +/- 0.3 | 7.3 +/- 0.2 | 9.0 +/- 0.5 | 8.1 +/- 0.3 |
| Prior surgery (%) | 15.4 | 15.4 | 11.3 | 11.5 |
| Length of Stay (median, IQR) | 5.7 (3.5, 10.1) | 3.9 (2.9, 5.4) | 5.8 (3.6, 9.8) | 4.0 (2.9, 5.7) |
| Matching Duration (median, IQR) | 4.2 (3.2, 5.9) | 3.9 (2.9, 5.4) | 4.4 (3.2, 6.0) | 4.0 (2.9, 5.7) |
| Time to Infection (median, IQR) | 7.9 (4.8, 16.1) | 0.0 (0.0, 0.0) | 8.0 (4.9, 16.6) | 0.0 (0.0, 0.0) |
| Prior antimicrobials (%) |  |  |  |  |
| β-lactams |  |  |  |  |
| Penicillins | 0.2 | 0.2 | 0.4 | 0.4 |
| Extended spectrum penicillins | 0.6 | 0.6 | 1.0 | 0.8 |
| Cephalosporins | 0.8 | 0.8 | 1.5 | 1.6 |
| Extended spectrum cephalosporins | 0.0 | 0.0 | 0.0 | 0.0 |
| Anti-Staphylococcal β-lactams | 0.0 | 0.0 | 0.0 | 0.0 |
| Other cell wall active agents |  |  |  |  |
| Glycopeptides | 0.0 | 0.0 | 0.0 | 0.0 |
| DNA synthesis inhibitors |  |  |  |  |
| Sulfonamides | 0.4 | 0.3 | 0.5 | 0.4 |
| Protein synthesis inhibitors |  |  |  |  |
| Fluoroquinolones | 0.5 | 0.4 | 1.5 | 1.1 |
| Tetracyclines | 0.5 | 0.3 | 0.6 | 0.6 |
| Macrolides | 0.2 | 0.2 | 0.8 | 0.8 |
| Lincosamides | 0.1 | 0.1 | 0.1 | 0.0 |
| Anti-anaerobic antibiotics |  |  |  |  |
| Metronidazole | 0.0 | 0.0 | 0.1 | 0.0 |
| Oral vancomycin | 0.0 | 0.0 | 0.1 | 0.0 |

**Table S7:** Baseline cohort characteristics for drug susceptible *P. aeruginosa*, and drug resistant *P. aeruginosa* cohorts. Abbreviations, SE, standard error; IQR, Interquartile Range.

| Feature | Drug susceptible *P. aeruginosa* |  | Drug resistant *P. aeruginosa* |  |
| --- | --- | --- | --- | --- |
|  | Case | Control | Case | Control |
| Sample size | 1,309 | 2,533 | 379 | 665 |
| Demographics |  |  |  |  |
| Age (mean, SE) | 68.3 +/- 0.5 | 68.4 +/- 0.4 | 69.3 +/- 0.8 | 69.6 +/- 0.6 |
| Gender (% female) | 49.7 | 51.4 | 44.1 | 47.3 |
| Elixhauser index (mean, SE) | 11.3 +/- 0.4 | 9.5 +/- 0.3 | 12.1 +/- 0.8 | 10.7 +/- 0.6 |
| Prior surgery (%) | 16.6 | 16.7 | 12.7 | 11.9 |
| Length of Stay (median, IQR) | 5.2 (3.7, 8.9) | 4.0 (2.9, 5.6) | 5.9 (3.7, 12.1) | 4.0 (3.0, 5.8) |
| Matching Duration (median, IQR) | 4.4 (3.4, 5.8) | 4.0 (2.9, 5.6) | 4.5 (3.3, 6.8) | 4.0 (3.0, 5.8) |
| Time to Infection (median, IQR) | 8.3 (5.0, 17.0) | 0.0 (0.0, 0.0) | 9.8 (5.1, 17.4) | 0.0 (0.0, 0.0) |
| Prior antimicrobials (%) |  |  |  |  |
| β-lactams |  |  |  |  |
| Penicillins | 0.1 | 0.1 | 0.3 | 0.3 |
| Extended spectrum penicillins | 1.0 | 0.9 | 0.3 | 0.3 |
| Cephalosporins | 2.1 | 2.4 | 1.3 | 0.9 |
| Extended spectrum cephalosporins | 0.2 | 0.2 | 0.0 | 0.0 |
| Anti-Staphylococcal β-lactams | 0.0 | 0.0 | 0.0 | 0.0 |
| Other cell wall active agents |  |  |  |  |
| Glycopeptides | 0.0 | 0.0 | 0.0 | 0.0 |
| DNA synthesis inhibitors |  |  |  |  |
| Sulfonamides | 1.0 | 0.9 | 1.6 | 1.1 |
| Protein synthesis inhibitors |  |  |  |  |
| Fluoroquinolones | 1.3 | 1.3 | 1.1 | 1.4 |
| Tetracyclines | 0.8 | 0.9 | 1.1 | 1.2 |
| Macrolides | 1.1 | 1.3 | 1.6 | 1.1 |
| Lincosamides | 0.2 | 0.2 | 0.3 | 0.2 |
| Anti-anaerobic antibiotics |  |  |  |  |
| Metronidazole | 0.0 | 0.0 | 0.0 | 0.0 |
| Oral vancomycin | 0.1 | 0.0 | 0.0 | 0.0 |

**Table S8:** Performance of conditional logistic regression and XGBoost models. Abbreviations, AUROC, area under the receiver operator curve; AUPRC, area under the precision-recall curve; PPV, positive predictive value; NPV, negative predictive value; LR+, likelihood ratio positive; LR-likelihood ratio negative.

| Model | AUROC (95% CI) | AUPRC (95% CI) | PPV | NPV | LR+ | LR- |
| --- | --- | --- | --- | --- | --- | --- |
| Conditional logistic regression |  |  |  |  |  |  |
| *E. coli* | 0.56 (0.55, 0.56) | 0.42 (0.42, 0.42) | 0.40 | 0.66 | 1.14 | 0.86 |
| ESBL *E. coli* | 0.59 (0.57, 0.61) | 0.43 (0.41, 0.45) | 0.40 | 0.71 | 1.26 | 0.77 |
| *K. pneumoniae* | 0.57 (0.56, 0.57) | 0.30 (0.30, 0.31) | 0.30 | 0.78 | 1.21 | 0.81 |
| ESBL *K. pneumoniae* | 0.66 (0.63, 0.69) | 0.58 (0.56, 0.61) | 0.51 | 0.68 | 1.49 | 0.65 |
| *C. difficile* | 0.66 (0.66, 0.66) | 0.50 (0.50, 0.51) | 0.47 | 0.74 | 1.52 | 0.61 |
| VS *E. faecalis* | 0.61 (0.61, 0.62) | 0.41 (0.40, 0.41) | 0.39 | 0.75 | 1.38 | 0.70 |
| VR *E. faecium* | 0.65 (0.64, 0.66) | 0.46 (0.44, 0.48) | 0.45 | 0.77 | 1.60 | 0.58 |
| MSSA | 0.63 (0.62, 0.63) | 0.42 (0.41, 0.43) | 0.39 | 0.77 | 1.40 | 0.67 |
| MRSA | 0.57 (0.56, 0.58) | 0.34 (0.34, 0.36) | 0.33 | 0.76 | 1.21 | 0.79 |
| DS *P. aeruginosa* | 0.57 (0.56, 0.58) | 0.39 (0.39, 0.40) | 0.37 | 0.71 | 1.20 | 0.82 |
| DR *P. aeruginosa* | 0.64 (0.62, 0.66) | 0.41 (0.39, 0.43) | 0.39 | 0.77 | 1.45 | 0.67 |
| XGBoost |  |  |  |  |  |  |
| *E. coli* | 0.57 (0.56, 0.58) | 0.43 (0.40, 0.45) | 0.40 | 0.68 | 1.13 | 0.82 |
| ESBL *E. coli* | 0.57 (0.55, 0.59) | 0.35 (0.25, 0.45) | 0.35 | 0.70 | 1.15 | 0.92 |
| *K. pneumoniae* | 0.56 (0.54, 0.58) | 0.38 (0.33, 0.42) | 0.40 | 0.69 | 1.29 | 0.87 |
| ESBL *K. pneumoniae* | 0.56 (0.49, 0.62) | 0.46 (0.18, 0.74) | 0.42 | 0.71 | 1.35 | 0.75 |
| *C. difficile* | 0.63 (0.60, 0.67) | 0.52 (0.46, 0.57) | 0.50 | 0.70 | 1.54 | 0.68 |
| VS *E. faecalis* | 0.59 (0.57, 0.61) | 0.46 (0.41, 0.50) | 0.43 | 0.70 | 1.33 | 0.75 |
| VR *E. faecium* | 0.59 (0.56, 0.61) | 0.47 (0.24, 0.70) | 0.50 | 0.62 | 1.35 | 0.83 |
| MSSA | 0.62 (0.60, 0.65) | 0.45 (0.38, 0.52) | 0.40 | 0.75 | 1.50 | 0.76 |
| MRSA | 0.57 (0.54, 0.60) | 0.37 (0.29, 0.45) | 0.33 | 0.69 | 1.05 | 0.97 |
| DS *P. aeruginosa* | 0.57 (0.55, 0.59) | 0.38 (0.33, 0.42) | 0.40 | 0.69 | 1.29 | 0.87 |
| DR *P. aeruginosa* | 0.62 (0.58, 0.67) | 0.49 (0.31, 0.67) | 0.41 | 0.61 | 1.06 | 0.97 |

**Table S9:**
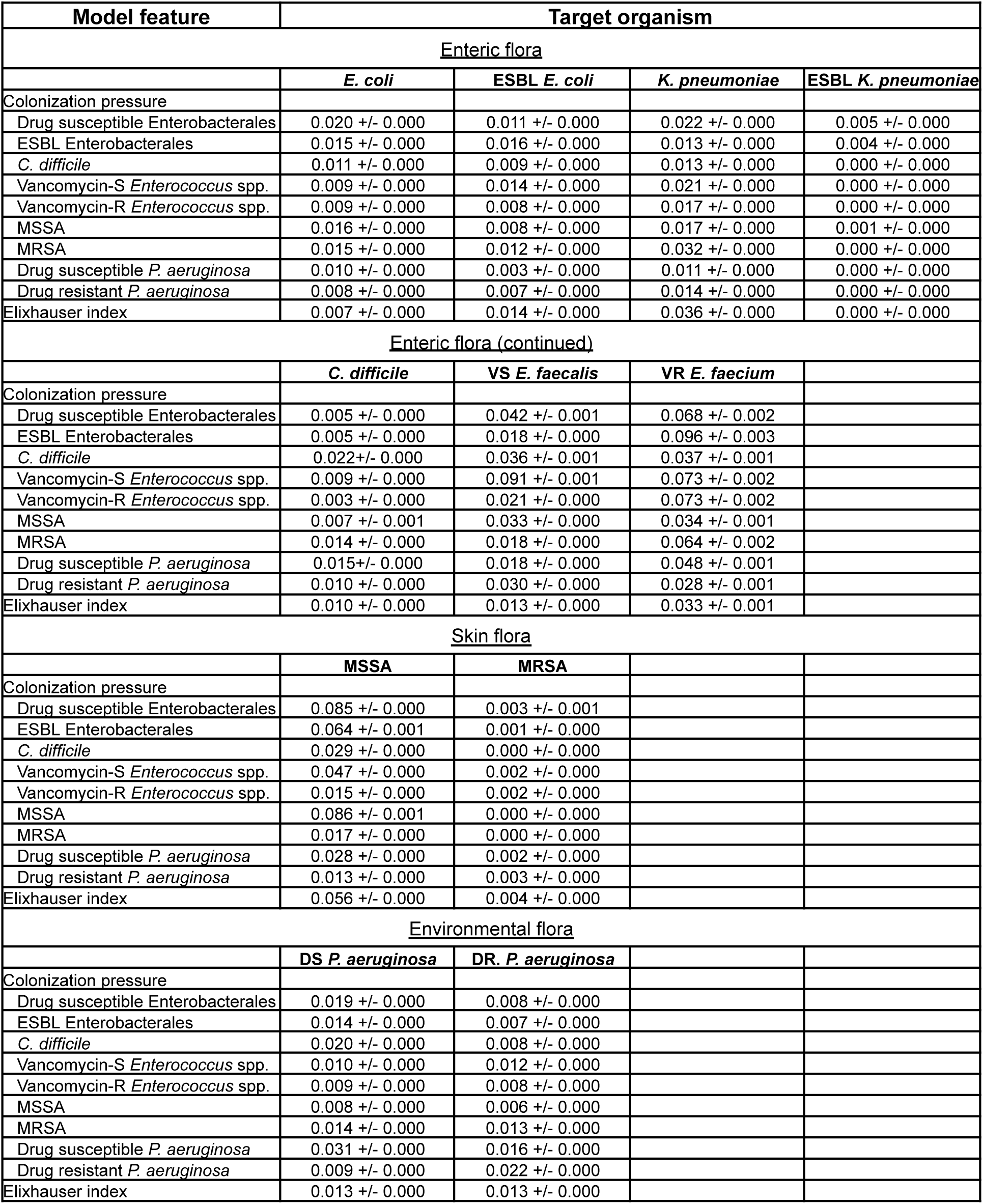
Mean SHAP values for XGBoost models.

## REFERENCES

1. CDC. Current HAI Progress Report | HAIs | CDC. 2024; published online Nov 25. https://www.cdc.gov/healthcare-associated-infections/php/data/progress-report.html (accessed June 9, 2025).

2. Warren BG, Turner NA, Addison R, et al. The Impact of Infection Versus Colonization on Clostridioides difficile Environmental Contamination in Hospitalized Patients With Diarrhea. Open Forum Infect Dis 2022; 9: ofac069.

3. Weinstein RA. Epidemiology and control of nosocomial infections in adult intensive care units. Am J Med 1991; 91: S179–84.

4. Weber DJ, Anderson D, Rutala WA. The role of the surface environment in healthcare-associated infections. Curr Opin Infect Dis 2013; 26: 338–44.

5. Martin RM, Cao J, Brisse S, et al. Molecular Epidemiology of Colonizing and Infecting Isolates of Klebsiella pneumoniae. mSphere 2016; 1: 10.1128/msphere.00261-16.

6. Eiff C von, Becker K, Machka K, Stammer H, Peters G. Nasal carriage as a source of Staphylococcus aureus bacteremia. Study Group. The New England journal of medicine 2001; 344: 11–6.

7. Worley J, Delaney ML, Cummins CK, DuBois A, Klompas M, Bry L. Genomic Determination of Relative Risks for Clostridioides difficile Infection From Asymptomatic Carriage in Intensive Care Unit Patients. Clin Infect Dis 2020. DOI:10.1093/cid/ciaa894.

8. Bonten MJ, Slaughter S, Ambergen AW, et al. The role of “colonization pressure” in the spread of vancomycin-resistant enterococci: an important infection control variable. Archives of internal medicine 1998; 158: 1127–32.

9. Ajao AO, Harris AD, Roghmann M-C, et al. Systematic review of measurement and adjustment for colonization pressure in studies of methicillin-resistant Staphylococcus aureus, vancomycin-resistant enterococci, and clostridium difficile acquisition. Infection control and hospital epidemiology 2011; 32: 481–9.

10. Masse J, Elkalioubie A, Blazejewski C, et al. Colonization pressure as a risk factor of ICU-acquired multidrug resistant bacteria: a prospective observational study. Eur J Clin Microbiol Infect Dis 2017; 36: 797–805.

11. Arvaniti K, Lathyris D, Ruimy R, et al. The importance of colonization pressure in multiresistant Acinetobacter baumannii acquisition in a Greek intensive care unit. Crit Care 2012; 16: R102.

12. Jolivet S, Lolom I, Bailly S, et al. Impact of colonization pressure on acquisition of extended-spectrum β-lactamase-producing Enterobacterales and meticillin-resistant Staphylococcus aureus in two intensive care units: a 19-year retrospective surveillance. J Hosp Infect 2020; 105: 10–6.

13. Merrer J, Santoli F, Vecchi CA-D, Tran B, Jonghe BD, Outin H. “Colonization Pressure” and Risk of Acquisition of Methicillin-Resistant Staphylococcus aureus in a Medical Intensive Care Unit. Infect Control Hosp Epidemiology 2000; 21: 718–23.

14. Rhee C, Kadri SS, Dekker JP, et al. Prevalence of Antibiotic-Resistant Pathogens in Culture-Proven Sepsis and Outcomes Associated With Inadequate and Broad-Spectrum Empiric Antibiotic Use. Jama Netw Open 2020; 3: e202899.

15. Ku JH, Tartof SY, Contreras R, et al. Antibiotic Resistance of Urinary Tract Infection Recurrences in a Large Integrated US Healthcare System. J Infect Dis 2024; 230: e1344–54.

16. Moore BJ, White S, Washington R, Coenen N, Elixhauser A. Identifying Increased Risk of Readmission and In-hospital Mortality Using Hospital Administrative Data. Méd Care 2017; 55: 698–705.

17. Chen T, Guestrin C. XGBoost: A Scalable Tree Boosting System. *arXiv* 2016. DOI:10.48550/arxiv.1603.02754.

18. Lundberg S, Lee S-I. A Unified Approach to Interpreting Model Predictions. arXiv 2017. DOI:10.48550/arxiv.1705.07874.

19. Wei Z, Sagers L, Kanjilal S. Source code for nosocomial pathogen acquisition due to hospital unit colonization pressure project. 2025; published online June 11. https://github.com/sanjatkanjilal/nosocomial-acquisition_colonization-pressure (accessed June 11, 2025).

20. Goldberger AL, Amaral LAN, Glass L, et al. PhysioBank, PhysioToolkit, and PhysioNet. Circulation 2000; 101: E215–20.

21. Klassert TE, Leistner R, Zubiria-Barrera C, et al. Bacterial colonization dynamics and antibiotic resistance gene dissemination in the hospital environment after first patient occupancy: a longitudinal metagenetic study. Microbiome 2021; 9: 169.

22. Chng KR, Li C, Bertrand D, et al. Cartography of opportunistic pathogens and antibiotic resistance genes in a tertiary hospital environment. Nat Med 2020; 26: 941–51.

23. Lax S, Sangwan N, Smith D, et al. Bacterial colonization and succession in a newly opened hospital. Sci Transl Med 2017; 9. DOI:10.1126/scitranslmed.aah6500.

24. Wolfensberger A, Clack L, Kuster SP, et al. Transfer of pathogens to and from patients, healthcare providers, and medical devices during care activity—a systematic review and meta-analysis. Infect Control Hosp Epidemiology 2018; 39: 1093–107.

25. Brooks B, Olm MR, Firek BA, et al. The developing premature infant gut microbiome is a major factor shaping the microbiome of neonatal intensive care unit rooms. Microbiome 2018; 6: 112.

26. Brooks B, Olm MR, Firek BA, et al. Strain-resolved analysis of hospital rooms and infants reveals overlap between the human and room microbiome. Nat Commun 2017; 8: 1814.

27. Puig-Asensio M, Diekema DJ, Boyken L, Clore GS, Salinas JL, Perencevich EN. Contamination of health-care workers’ hands with Escherichia coli and Klebsiella species after routine patient care: a prospective observational study. Clin Microbiol Infect 2020; 26: 760–6.

28. Rodríguez-Acelas AL, Almeida M de A, Engelman B, Cañon-Montañez W. Risk factors for health care–associated infection in hospitalized adults: Systematic review and meta-analysis. Am J Infect Control 2017; 45: e149–56.

29. Bonten MJM. Colonization pressure: a critical parameter in the epidemiology of antibiotic-resistant bacteria. Critical care (London, England) 2012; 16: 142.

30. Ajao AO, Johnson JK, Harris AD, et al. Risk of Acquiring Extended-Spectrum β-Lactamase–Producing Klebsiella Species and Escherichia coli from Prior Room Occupants in the Intensive Care Unit. Infect Control Hosp Epidemiology 2013; 34: 453–8.

31. Nseir S, Blazejewski C, Lubret R, Wallet F, Courcol R, Durocher A. Risk of acquiring multidrug-resistant Gram-negative bacilli from prior room occupants in the intensive care unit. Clin Microbiol Infect 2011; 17: 1201–8.

32. Drees M, Snydman DR, Schmid CH, et al. Prior Environmental Contamination Increases the Risk of Acquisition of Vancomycin-Resistant Enterococci. Clin Infect Dis 2008; 46: 678–85.

33. Willems RJL, Hanage WP, Bessen DE, Feil EJ. Population biology of Gram-positive pathogens: high-risk clones for dissemination of antibiotic resistance. FEMS microbiology reviews 2011; 35: 872–900.

34. McDonald EG, Dendukuri N, Frenette C, Lee TC. Time-Series Analysis of Health Care–Associated Infections in a New Hospital With All Private Rooms. JAMA Intern Med 2019; 179: 1501–6.

35. Liu CM, Price LB, Hungate BA, et al. Staphylococcus aureus and the ecology of the nasal microbiome. Science advances 2015; 1: e1400216–e1400216.

36. Zmora N, Zilberman-Schapira G, Suez J, et al. Personalized Gut Mucosal Colonization Resistance to Empiric Probiotics Is Associated with Unique Host and Microbiome Features. Cell 2018; 174: 1388–1405.e21.

37. Bogaert D, Belkum A van, Sluijter M, et al. Colonisation by Streptococcus pneumoniae and Staphylococcus aureus in healthy children. Lancet 2004; 363: 1871–2.

38. Zeise KD, Woods RJ, Huffnagle GB. Interplay between Candida albicans and Lactic Acid Bacteria in the Gastrointestinal Tract: Impact on Colonization Resistance, Microbial Carriage, Opportunistic Infection, and Host Immunity. Clin Microbiol Rev 2021; 34: e00323–20.

39. González LM, Mukhitov N, Voigt CA. Resilient living materials built by printing bacterial spores. Nat Chem Biol 2020; 16: 126–33.

